# Altered frontolimbic activity during virtual reality-based contextual fear learning in patients with posttraumatic stress disorder

**DOI:** 10.1101/2022.06.07.22275758

**Authors:** Sebastian Siehl, Manon Wicking, Sebastian Pohlack, Tobias Winkelmann, Francesca Zidda, Frauke Steiger-White, Frauke Nees, Herta Flor

## Abstract

**Background:** Deficiency in contextual and enhanced responding in cued fear learning may contribute to the development of posttraumatic stress disorder (PTSD). We examined the responses to aversive Pavlovian conditioning with an unpredictable spatial context as conditioned stimulus compared to a predictable context. We hypothesized that the PTSD group would depict less hippocampal and ventromedial prefrontal cortex (vmPFC) activation during acquisition and extinction of unpredictable contexts and an overreactive amygdala response in the predictable contexts compared to controls.

**Methods:** A novel combined differential cue-context conditioning paradigm was applied using virtual reality with spatial contexts that required configural as well as cue processing. We assessed 20 patients with PTSD, 21 healthy trauma-exposed (TC) and 22 non-trauma-exposed (HC) participants using functional magnetic resonance imaging (fMRI), skin conductance responses and self-report measures.

**Results:** During fear acquisition patients with PTSD compared to TC showed lower activity in the hippocampi in the unpredictable and higher activity in the amygdalae in the predictable context. During fear extinction, patients compared to TC showed lower brain activity in the vmPFC in the predictable context. There were no significant differences in self-report or skin conductance responses among the groups.

**Conclusions:** Our results suggest that patients with PTSD differ in brain activation from controls in regions such as the hippocampus, the amygdala and the vmPFC in the processing of unpredictable and predictable contexts. Deficient encoding of more complex configurations might lead to a preponderance of cue-based predictions in PTSD. Exposure-based treatments need to focus on improving predictability of contextual processing and reducing enhanced cue reactivity.

## Introduction

Studies of fear learning, using pavlovian conditioning (Pavlov, 1927), have greatly advanced our understanding of the psychobiological mechanisms of posttraumatic stress disorder (PTSD), which is characterized by symptoms like re-experiencing, avoidance, hyperarousal and alterations in mood and cognition (American Psychiatric Association, 2013). In pavlovian fear conditioning, an originally neutral cue or context is paired with a biologically relevant stimulus (unconditioned stimulus (US)), to become a conditioned stimulus (CS, cue or context). This CS can then elicit a conditioned response that is similar to the response to the unconditioned response (UR), without the US being present. This association between the US and the CS is learned during fear acquisition and can be overwritten during fear extinction. Deficient context learning has been at the center of recent psychobiological models of posttraumatic stress disorder (PTSD; Liberzon & Abelson, 2016; Maren et al., 2013; Shalev et al., 2017) and has been associated with typical PTSD symptoms (Shalev et al., 2017). Spatial contexts have most often been examined in contextual learning and it has been proposed that patients with PTSD may not be able to sufficiently discriminate safe from dangerous contexts and thus maintain a fear response even in safe contexts (Acheson et al., 2012; Flor & Wessa, 2010). Deficient context learning has been associated with lower functional activity in the hippocampus and ventromedial prefrontal cortex (vmPFC; Acheson et al., 2012; Garfinkel et al., 2014; Pitman et al., 2012). In addition, enhanced learning of trauma-related cues has been observed in patients with PTSD, which was associated with higher functional activity in the amygdala (Garfinkel et al., 2014; Pitman et al., 2012). A variety of study protocols were developed to investigate context conditioning (Lonsdorf et al., 2017) using virtual reality in human subjects (Glenn et al., 2017; Kroes et al., 2017). Two factors that are conducive to an environment being perceived as a context are its longer representation time and its higher complexity compared to single cues (Lonsdorf et al., 2017). The larger the time frame, in which a US can occur, the larger the unpredictability and the higher the levels of anxiety (Indovina et al., 2011; Schmitz & Grillon, 2012). With the US being presented within a longer presentation phase of a given context it cannot be directly associated with an object within the context, which is why contexts are usually considered to be unpredictable of the US. However, a given context might be recognizable by a single cue, such as a table, which is present in one but not in a second context. Previous context conditioning studies have shown that patients with PTSD show difficulties in contextual fear acquisition, but improve when cues are added to predict if a context is dangerous or safe (Steiger et al., 2015). Furthermore, patients with PTSD show reduced capacity to use context information to regulate fear responses (Garfinkel et al., 2014) during fear extinction (Rougemont-Bücking et al., 2011), memory of the extinction (extinction recall; Milad et al., 2009) and when the already extinguished fear is returning (fear renewal; Wicking et al., 2016). Here, contexts were defined as steady images of an office (Garfinkel et al., 2014; Rougemont-Bücking et al., 2011) or virtual reality scenes of different rooms (Wicking et al., 2016), in which contexts could be distinguished by retrieving a single object from the environments, making it a predictable context. In configural learning (Acheson et al., 2012; Rudy et al., 2004; Rudy & O’Reilly, 1999), multiple objects that are associated to each other form a conjunctive representation of a context. Acheson et al. (2012) hypothesized that lower hippocampal activity leads to an inability of configural-based learning in patients with PTSD. Instead, cue-based associations will be formed, in which a single cue is assumed to predict the occurrence of the US, independent of its predictability or the complexity of the surrounding information (Acheson et al., 2012; Lonsdorf et al., 2017). Each individual cue is then potentially able to elicit a fear response across contexts, with the amygdala being more active (Phillips & Ledoux, 1992), independent of the context being safe or dangerous.

The aim of our study was to compare an unpredictable context based on configural learning with a predictable context, in which a single cue predicted the occurrence of a US within a context in patients with PTSD and trauma-naïve and trauma-experienced controls. To this end we developed a new VR-based, combined cue-context conditioning paradigm. This new approach defined contexts via a configural learning approach, in which the configuration of furniture in a room defined a particular context. Participants were presented with unpredictable and predictable contexts as CS during fear acquisition and extinction. We hypothesized that patients with PTSD in comparison to HC and TC subjects would a) show lower BOLD activity in the hippocampi and vmPFCs, b) show lower skin conductance response c) and report higher arousal, valence and contingency ratings during acquisition of the unpredictable context. In the predictable context, we expected higher BOLD activity in the amygdalae in patients with PTSD in comparison to TC and HC subjects. During contextual fear extinction, we hypothesized higher BOLD activity in the amygdalae and lower activity in the vmPFCs for patients with PTSD in comparison to both control groups in both contexts.

## Methods and Materials

### 2.1 Participants

Twenty patients suffering from PTSD, 21 age- and sex matched TC and 22 HC subjects participated in this study (*Table 1*; for details on recruitment and inclusion criteria see *Suppl. methods*). The study was carried out in accordance with the Code of Ethics of the World Medical Association (Declaration of Helsinki, 2013) and was approved by the Ethical Review Board of the Medical Faculty Mannheim, Heidelberg University. A all participants gave written informed consent.

**Table 1.**
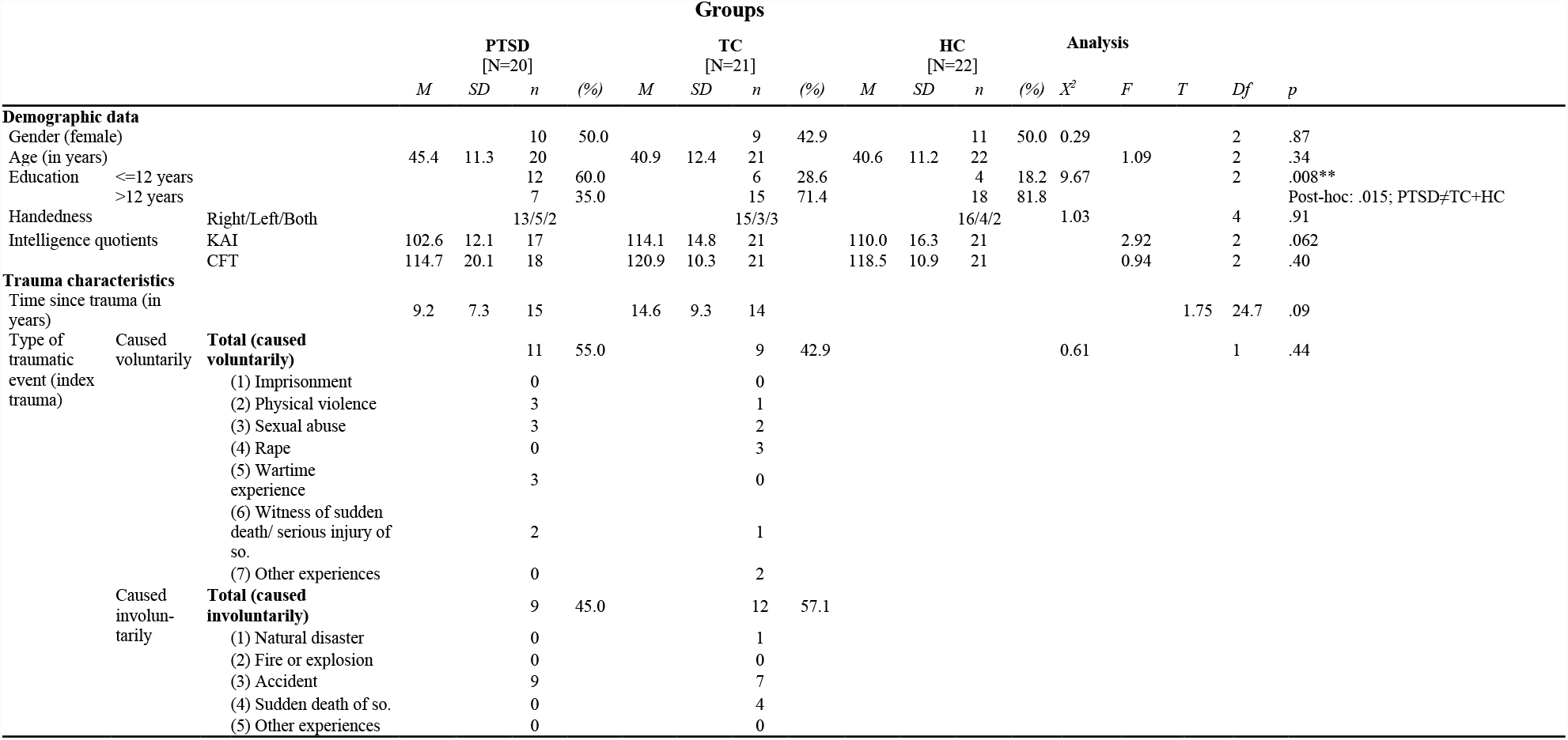
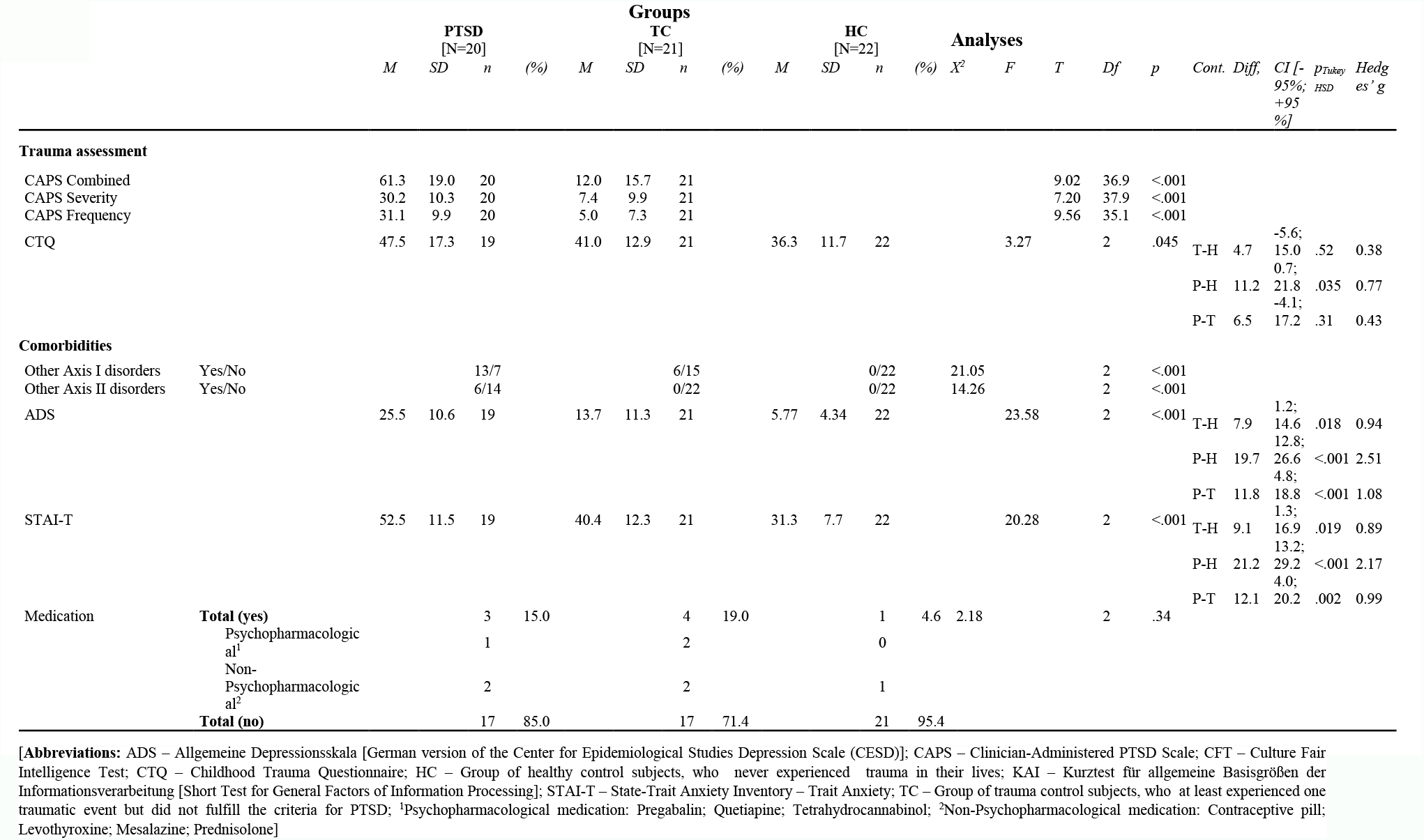
Demographic and clinical Characteristics of study sample.

### 2.2 Procedure and study design

The study consisted of two assessments on two consecutive days, each lasting for approximately five hours. On the first day, participants completed questionnaires and clinical assessments on PTSD and participated in the Structured Clinical Interviews (SCID I+II; Fydrich et al., 1997; Wittchen et al., 1997). During the first experimental phase participants completed a training- and habituation phase outside the Magnetic Resonance Imaging (MRI) scanner, while sitting in front of a computer screen with a head mounted display (HMD). Participants then determined the intensity of the painful stimulus that served as US (*Suppl. methods*) before completing the context and cue acquisition phases inside the MRI scanner. On the second day, participants took part in the context and cue extinction phases inside the MRI scanner. This was followed by a final testing phase including cognitive and neuropsychological assessments.

### 2.3 Stimuli and experimental procedure

During the experimental phase, participants were passively navigated through virtual contexts (living rooms) on a parabola shaped trajectory with a constant slow-paced walking speed of 0.45 km/h and an egocentric viewpoint (*Figure 1*). A total of four different contexts and two different cues (CS+, CS-, coloured sqares) were presented during acquisition and extinction (*Figure 1* and see *Suppl. Methods)* in a differential conditioning paradigm, where the CS+ signaled the US and the CS-its absence. The US was a painful stimulus. In one context the cue did not predict the US (unpredictable [unpred]), in the other context a cue reliably predicted the US (predictable [pred]). There were two additional contexts with different configuration of the furniture in which no painful stimulus occurred [safe]. The perspective rotated slightly from right to left and right again, so that each of the four walls of a room was entirely visible at least once. The virtual contexts consisted of several objects (bookshelves, chest of drawers, floor lamp, potted plant, racks, seating corner, television, *Figure 1*) and were built using an online software toolbox Open-Source Graphics Rendering Engine (OGRE; http://ogre3d.org) and the support of a software company (Glodeck Software GmbH) using Visual Studio Professional (2010, Redmond, WA, USA). The arrangement of the objects differed for each context but the objects were identical, thus forcing configural processing for context differentiation. Two colored CS+/-squares were presented on the walls of each context in a counterbalanced fashion.

**Figure 1.**
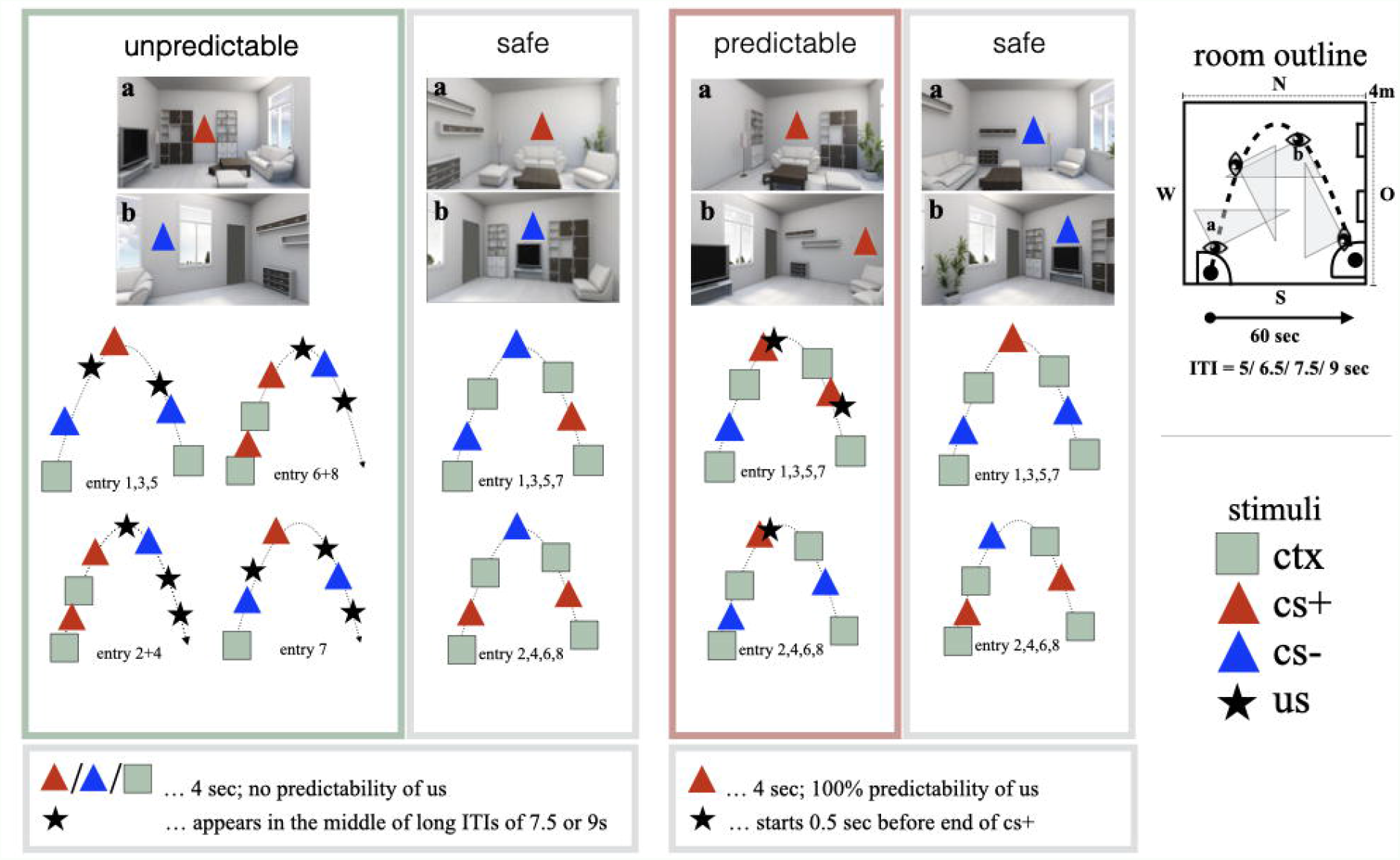
This figure depicts the four conditions (unpredictable, predictable, 2x safe) exemplary for both acquisition phases. Abbreviations: cs - conditioned stimulus; ctx - context; ITI - Inter-Trial-Interval; us - unconditioned stimulus]

### 2.4 Statistical analysis

All statistical analyses were performed in R-Statistics (Team, 2013). Data were assessed for outliers, normal distribution, homoscedasticity and multicollinearity. All assumptions were met, if not mentioned otherwise below. Demographic and clinical data as well as self reports and SCRs were analyzed with analyses of variance (ANOVAs) or independent t-tests in case of two sample comparisons (e.g. trauma characteristics). Chi-square tests were performed to assess statistical differences in frequency distributions (e.g. gender). For the ROI analyses we performed ANOVAs including the factors group (PTSD, TC, HC) x context (unpred, pred) x hemisphere (left, right) separately for the ACQ and EXT phase as well as brain region (hippocampus, amygdala, vmPFC). Based on our a priori hypotheses, we only compared the functional activity within the above mentioned ROIs and in the contexts, in which a painful stimulus occurred (unpred, pred). We applied Bonferroni corrections to counteract Type 1 errors due to multiple comparisons. We further applied Tukey’s honestly significant difference (Tukey’s HSD) test as post-hoc single-step comparison procedure. There were few cases of missing data, motion artefacts or outliers (see *Results*).

The *supplementary methods* include information about participants, stimuli and experimental procedure, skin conductance responses (SCR), clinical and neuropsychological assessments, self-reports, MRI data acquisition and analysis, manipulation check and statistical analysis.

## 3 Results

### 3.1 Sample characteristics

The experimental groups did not significantly differ in any of the demographic variables except for education (*X*^*2*^(2, 62) = 9.67, *p* = .008). Patients with PTSD had a significantly lower level of education than the TC and HC group (*p* = .015; *Table 1*). All detailed information on demographic data, trauma severity, PTSD assessment and comorbidities can be found in *Table 1* and the *Suppl. Results*. In addition, we describe more detailed results on personality traits and neuropsychological assessment in *Suppl. Table 1* and in the *Suppl. Results*. There was no significant difference between the experimental groups on any of the debriefing questions concerning the difficulty of the study (*Suppl. Table 2, Suppl. Results*).

### 3.2 Functional magnetic resonance imaging

During acquisition, there was a significant interaction of group x context (*F*_*group x context*_ (2, 47) = 3.42, *p* = .04) in the hippocampus with significantly higher beta values in HCs than patients with PTSD in the unpredictable context (p_adj._ = .035). In addition, HCs had significantly higher beta values in the hippocampus during the unpredictable in comparison to the predictable context (p_adj._ = .0001). For the interaction group x context (*F*_*group x context*_ (2, 47) = 3.42, *p* = .04) in the amygdala a post-hoc t-test revealed a significantly higher beta value in the predictable context in HCs than in patients with PTSD (p_adj._ = .006) and HCs in comparison to TCs (p_adj._ = .02). Furthermore, HCs had significantly higher beta values in the unpredictable in comparison to the predictable context (p_adj._ = .003). There was no other significant main effect or interaction during acquisition (*Figure 2; Table 2a*).

**Table 2a.**
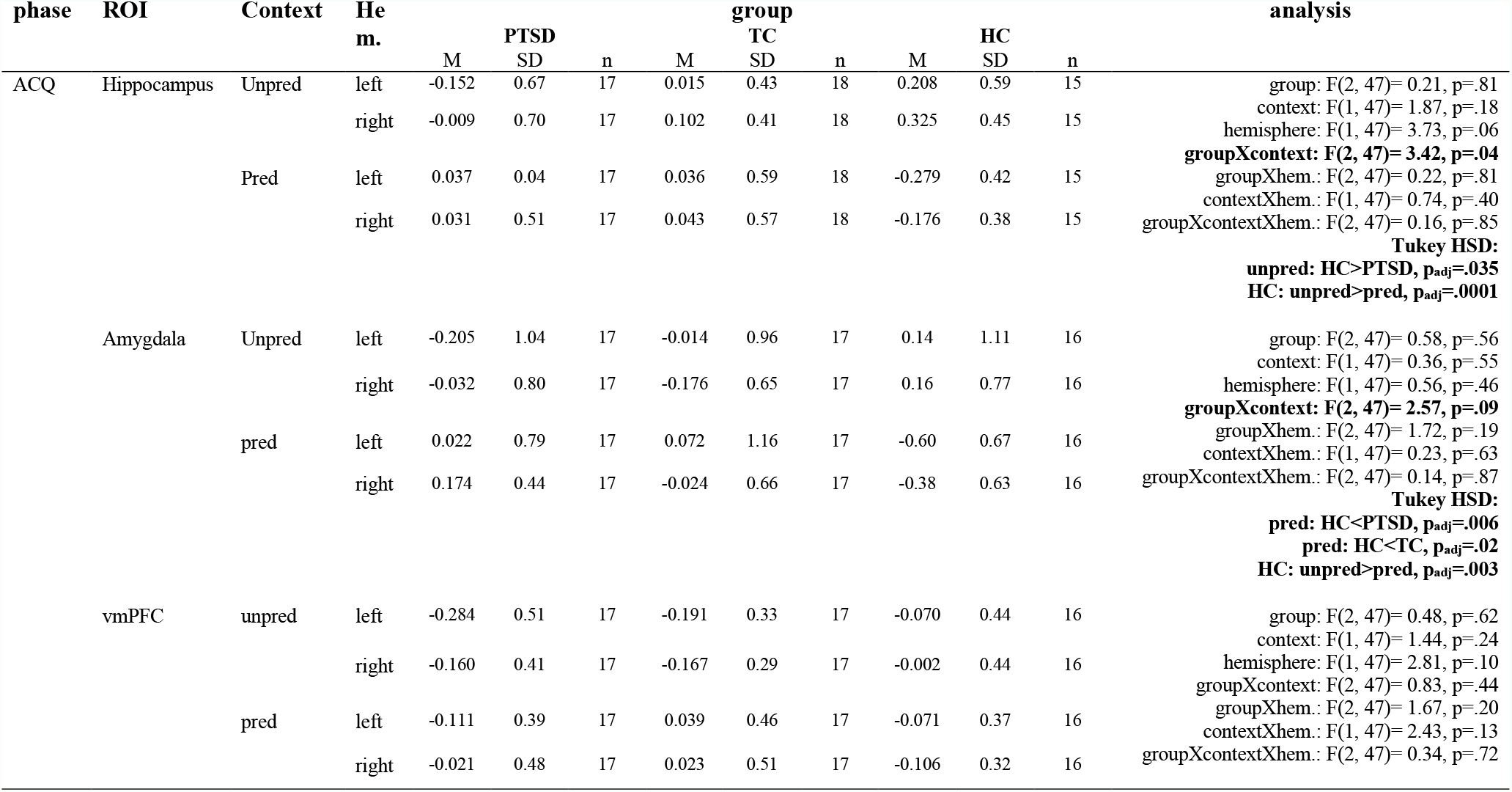
Extracted beta values for Region-of-Interest (ROI) analyses on the Hippocampi, Amygdalae and vmPFC for context unpredictable (unpred) and context predictable (pred) during acquisition and for each group (HC, PTSD, TC).

**Figure 2.**
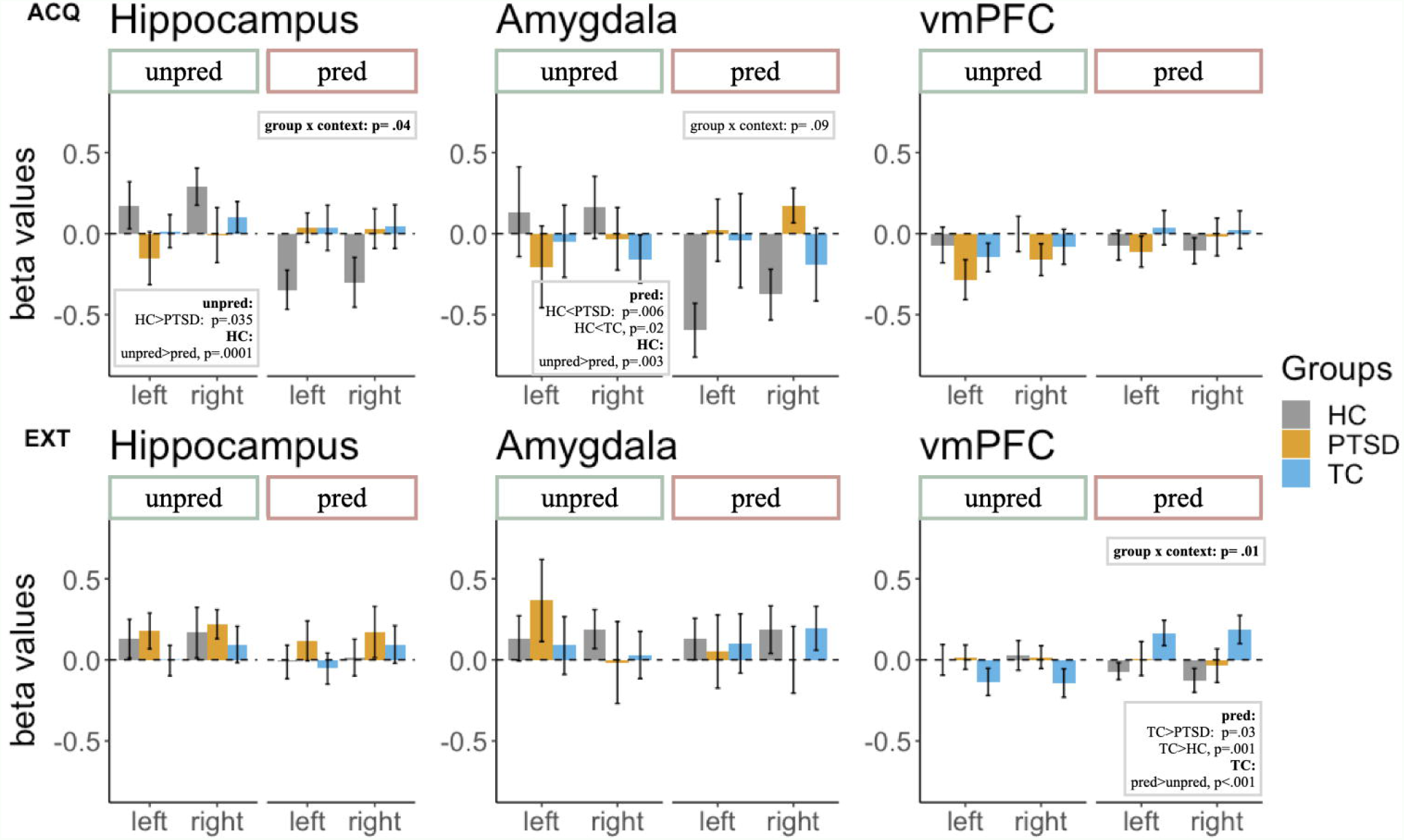
Extracted beta values for region-of-interest (ROI) analyses on the hippocampi, amygdalae and vmPFC during acquisition (ACQ; top row) and extinction (EXT; bottom row) for each group (HC, PTSD, TC). [Abbreviations: HC - Healthy control subjects without trauma experience; pred - predictable; PTSD - patients with PTSD; TC - healthy control subjects with trauma experience; unpred - unpredictable; vmPFC - ventromedial prefrontal cortex]

During extinction, we found a significant interaction of group x context (*F*_*group x context*_ (2, 46) = 5.03, *p* = .01) in the vmPFC. A post-hoc t-test revealed a significantly higher beta value in TCs than patients with PTSD (p_adj._ = .03) and TCs in comparison to HCs (p_adj._ = .001), both in the predictable context. Furthermore, TCs showed higher beta values in the vmPFC in the predictable than in the unpredictable context (p_adj._ < .001). There was no other significant main effect or interaction during extinction (*Figure 2; Table 2b*).

**Table 2b.**
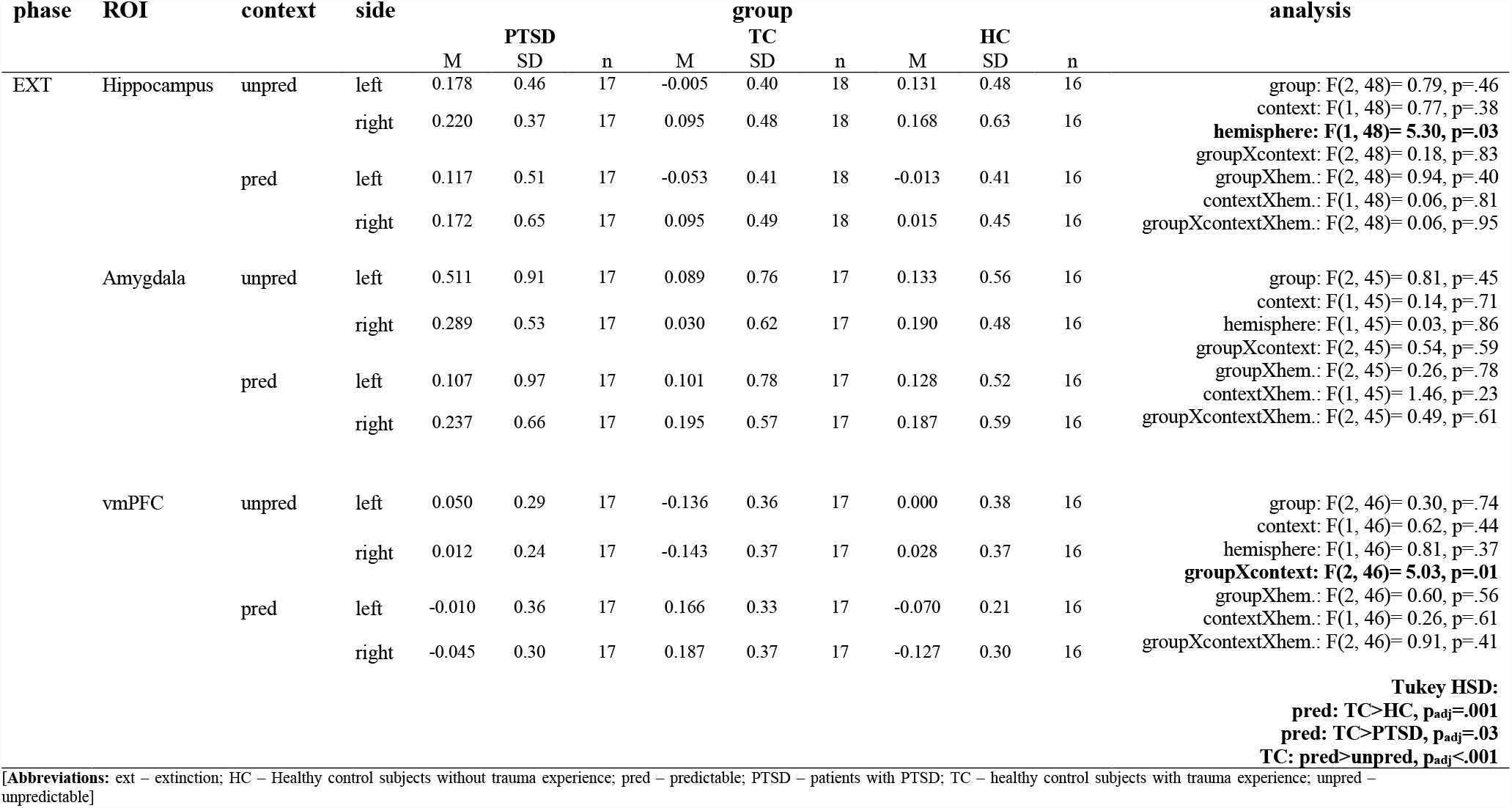
Extracted beta values for Region-of-Interest (ROI) analyses on the Hippocampi, Amygdalae and vmPFC for context unpredictable (unpred) and context predictable (pred) during extinction and for each group (HC, PTSD, TC).

### 3.3 Skin Conductance

During acquisition, there was a significant main effect for context in the unpredictable (*F*_*context*_(1, 36) = 14.55, *p* < .001) in comparison to the safe and the predictable (*F*_*context*_(1, 34) = 66.07, *p* < .001) in comparison to the safe contexts. There was also a main effect of group in the predictable condition (*F*_*group*_(2, 34) = 5.45, *p* < .009) with higher SCRs for participants in the HC group than patients with PTSD or TCs. We did not find a significant main effect of group during the unpredictable context condition, nor any significant interaction of group x context (*Figure 3; Suppl. Table 4a*). Patients with PTSD in comparison to HC and TC subjects showed a similar SCR in the unpredictable context. However, we found a significantly lower SCR in the predictable context for patients with PTSD in comparison to the two healthy control groups during the predictable context.

**Figure 3.**
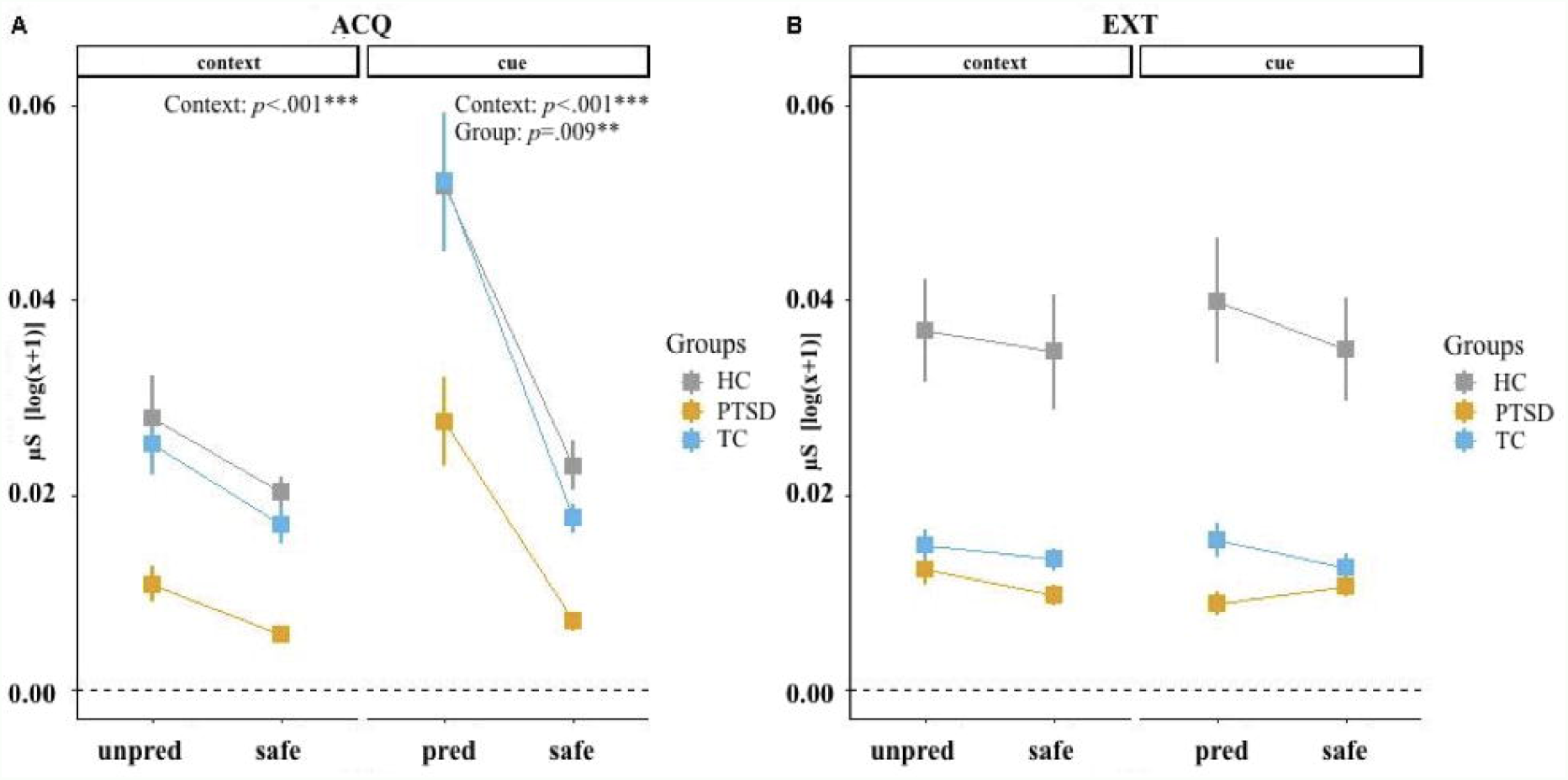
SCRs across each of the four conditions (unpred, pred, 2xsafe), two phases (ACQ, EXT) and each group (HC, PTSD, TC). A) ACQ phase. B) EXT phase. [Abbreviations: ACQ - Acquisition; CTX - Context; EXT - Extinction; HC - Healthy control subjects without trauma experience; pred - Predictable; PTSD - patients with PTSD; SCR - Skin conductance response; TC - healthy control subjects with trauma experience; unpred - Unpredictable]

Differences in SCRs between CS+ - CS-. During contextual fear acquisition, we found a significant main effect of group for the mean difference of CS+-CS-in the unpredictable (*F*_*group*_(2, 36) = 3.93, *p* = .029) in comparison to the safe and in the predictable (*F*_*context*_(1, 34) = 62.59, *p* < .001) in comparison to the safe contexts. Patients with PTSD and TC subjects showed a significantly lower CS+/-differentiation than the HC subjects in the unpredictable contexts. In the predictable contexts, the groups did not significantly differ in their difference scores but showed overall higher scores in the predictable context than in the safe context. There was no other significant main effect or interaction of group x context. During context extinction, we found a significant main effect of context for the mean difference of CS+/CS-(*F*_*context*_(1, 35) = 5.47, *p* = .025) with lower SCRs in the unpred in comparison to the safe context. There was no other significant main effect of interaction of group x context (*Suppl Figure 3; Suppl. Table*

### 3.4 Self-report

Ratings across contexts. We found significant main effects of phase (HAB, ACQ, EXT), separately across all four contexts (unpred, pred, 2x safe) for the arousal and contingency ratings with the highest scores during acquisition and the lowest scores during extinction. For the valence ratings, we found a significant main effect of phase in the unpredictable context, with higher ratings in the acquisition than in the habituation or extinction phase and a significant group x phase interaction (*Suppl. Figure 2a; Suppl. Table 3b-d*). Patients with PTSD in comparison to HC and TC subjects reported similar arousal, valence and contingency ratings during the unpredictable context. There was no significant difference in the ratings between the groups in the predictable context.

Differences in ratings between CS+ and CS-. There were significant main effects of phase during acquisition for ratings of arousal (*F*_*phase*_(1, 56) = 39.13, *p* < .001), valence (*F*_*phase*_(1, 56) = 21.54, *p* < .001) and contingency (*F*_*phase*_(1, 56) = 42.89, *p* < .001), showing that all three groups successfully learned the differential cue conditioning in the acquisition phase. There were no other significant main effects of phase or group nor any significant interaction of group x phase (*Suppl. Figure 2b*; *Suppl. Table 3e*).

## Discussion

The present study investigated differences in functional activity in the hippocampus, amygdala and vmPFC in unpredictable and predictable contextual fear learning in patients with PTSD in comparison to healthy trauma or non-trauma exposed control subjects. During contextual fear acquisition, patients with PTSD showed significantly lower functional brain activity in the hippocampi in the unpredictable context and significantly higher activity in the amygdalae and SCR in the predictable context than HC subjects. In addition, HC subjects displayed a main effect of context, with higher activity in the unpredictable in comparison to the predictable context for the hippocampus and amygdala. During contextual fear extinction, patients with PTSD showed significantly lower activity in the vmPFC in the predictable context than TC subjects. Here, TC subjects had significantly higher activity patterns in the vmPFC in the unpredictable in comparison to the predictable context. There were no significant differences between the groups in the behavioral ratings for the contexts. Learning about the predictability of an aversive stimulus within a given context and overwriting what one has learned during extinction are key mechanisms of associative learning. In this novel VR-based combined context-cue conditioning paradigm, we show that both context and cue in context conditioning are altered in patients with PTSD.

During contextual fear acquisition, we showed that patients with PTSD display lower functional brain activity in the hippocampi in the unpredictable and higher functional brain activity in the amygdalae in the predictable context. This fits well with findings that the hippocampi are involved in contextual fear acquisition of complex environments (Maren et al., 2013) and that difficulties in contextual fear acquisition in patients with PTSD might be associated with difficulties in configural learning, which was required in our study (Acheson et al., 2012). The amygdala in contrast has been associated to cued processing (Maren et al., 2013; Rudy, 2009) and is well established as a region of interest in PTSD within a larger brain network processing salience and threat (Shalev et al., 2017). Previous studies have shown that behavioural and psychophysiological responses as well as functional brain activity during contextual fear learning often show a contrasting picture (Baeuchl et al., 2015; Steiger et al., 2015; Wicking et al., 2016). In line with these findings, we did not observe any differences in behavioural valence, arousal or contingency ratings between the experimental groups, neither during acquisition nor extinction. We showed significantly lower SCRs in patients with PTSD in comparison to both healthy control groups as a response to the predictable context. This is in line with findings in cue-only conditioning studies reporting higher amygdala activity in patients with PTSD suggesting increased threat detection with lower peripheral psychophysiological reactions. In line with this, we did not find a significant correlation between amygdala activity and SCRs. This might be interpreted in a way that higher amygdalae activity in patients suggests that some information about the threat is learned and increases the predictability the cue, which might minimize the peri-physiological response. Our study highlights that it is essential to distinguish the predictive power of the occurrence of an aversive stimulus in a given context and whether a threat can be predicted by a single cue or only by a more complex configuration of objects (e.g. furniture). Furthermore, operationalizing context learning via a configural learning approach facilitates to overserve cue and context related responses in the same environment.

During contextual fear extinction, we showed higher functional brain activity in the vmPFC in TC subjects in comparison to patients with PTSD and HC subjects. The vmPFC is commonly found to be engaged during fear extinction (Lang et al., 2009; Maren et al., 2013), and has been repeatedly associated with the inhibition of fear responses. Difficulties in contextual fear extinction and extinction recall in PTSD have been associated with lower functional activity in the vmPFC (Glenn et al., 2017; Wicking et al., 2016), which is also related to structural white and gray matter reduction in the vmPFC (Siehl et al., 2018, 2020). While some studies do report hippocampal involvement during contextual fear extinction (Milad et al., 2007) or contextual retrieval (Maren et al., 2013), the findings in the literature do not yield a consistent picture. We suggest that one common factor between contextual fear acquisition and extinction might be the difference in functional brain activation between unpredictable and predictable contexts in healthy control subjects but not in patients with PTSD. Our findings are in line with recent models examining PTSD in a predictive-coding framework (Seriès, 2019). Forming accurate predictions about the safety of one’s environment is necessary for the ability to plan actions and interpret bodily sensations. If uncertainty about the danger of safety of the surrounding environment is high, due to difficulties in forming a stable mental representation of the context through configural learning, the prediction may favor safety. High levels of anxiety might be the consequence for patients with PTSD, preparing for a potential danger and paying a metabolic price for a contextual prediction that fails to be updated. Future studies will have to further assess the role of building (via configural learning) a mental representation of a context in contrast to elemental contextual learning as two important factors for predictability in contextual fear learning.

### 4.2 Limitations

Two main limitations apply to our study. Whereas the complexity of the design allows for the simultaneous examination of context- and cue-related triggers and their interaction, the design limits the choice of where to select the context triggers in each environment. To minimize overlapping, or additive effects in SCR or BOLD activity, the triggers had to be far enough apart from each other. This, however, extended each trial to 50 secs, which in turn limited our total number of trials per given condition to eight. With this rather low number of trials, each missing data point became a potential dropout. The interdependence of the triggers in the unpredictable context was also a problem. Here, almost each trial represented a unique composition of positions for the CSs, US and context triggers. A fear response in a given context is most likely not limited to a single predictive cue or multiple contextual features but might also generalize to objects being non-predictive to the occurrence of the aversive stimulus. This is, however, a problem in many studies and not limited to our study (Lonsdorf et al., 2017) and the presented results have to be interpreted within the boundaries of the study design. A second limitation concerns a rather high number of potential non-responders in the SCR assessment. The SCR was measured on the foot, instead of the hand of participants, because participants received the painful stimulus on the left hand and responded with the response pad on the right hand. The signal on the foot might not have been strong enough. A potential solution could be to measure SCR on the shoulder instead (van Dooren et al., 2012).

### 4.3 Conclusions

In this cross-sectional study, we show that patients with PTSD when compared to TC and HC subjects display lower hippocampal activation in unpredictable and higher amygdala activity in predictable contextual fear acquisition as well as lower activity in the vmPFC during extinction of predictable contexts. Our results support the model that patients with PTSD show deficiencies in configural learning, while being more sensitive to cue-based learning and highlight the importance of predictability of fear in contextual learning. Trauma-focused exposure-based treatments might benefit from increasing predictability during the integration of contextual features within traumatic memories during exposure.

## Data Availability

We report how we determined our sample size, all data exclusions, all inclusion/exclusion criteria, whether inclusion/exclusion criteria were established prior to data analysis, all manipulations, and all measures in the study. Ethical restrictions to protect participant confidentiality prevent us from making study data publicly available. This also refers to the analysis/experimental code, and any other digital materials, where participant-related information (like sex or psychopathological status) are also included. Readers seeking access to the study data and materials should contact the corresponding author based on a formal collaboration agreement. This formal collaboration agreement indicates that data will be shared with other researchers who agree to work with the authors, and for the sole purpose of verifying the claims in the paper. The data and materials will be released to requestors after approval of this formal collaboration agreement by the local Ethics Committee of the Medical Faculty Mannheim.

## Disclosures

The authors report no financial relationships with commercial interests.

## Acknowledgements

We thank Birgül Sarun and Claudia Stief for help in data acquisition and Michaela Ruttorf for advice regarding data analysis. This work was supported by a grant of the Deutsche Forschungsgemeinschaft (SFB636/C1 to H.F).

## Notes

**Author Note:** All authors declare that the research was conducted in the absence of any commercial or financial relationships that could be construed as a potential conflict of interest. The datasets and scripts for analyses are made available online. Readers seeking further information are welcome to contact the corresponding author.

### Competing Interest Statement

The authors have declared no competing interest.

### Author Declarations

The study was carried out in accordance with the Code of Ethics of the World Medical Association (Declaration of Helsinki, 2013) and was approved by the Ethical Review Board of the Medical Faculty Mannheim, Heidelberg University. A all participants gave written informed consent.

