## Supplementary material for "Altered frontolimbic activity during virtual reality-based contextual fear learning in patients with posttraumatic stress disorder": Suppl. Text

### Supplemental Information

### Supplemental methods

#### *Participants*

We recruited participants through the outpatient clinic of the Institute of Cognitive and Clinical Neuroscience and advertisement on the recruitment website of the Central Institute of Mental Health in Mannheim. In addition, we recruited patients from local psychotherapy and psychiatry practices as well as local clinics and outpatient units. Prior to testing, a telephone screening was conducted with all participants. The following exclusion criteria were applied: any traumatic experience before the age of 18 years, borderline personality disorder, comorbid current or lifetime psychotic symptoms, current substance dependence or abuse, cardiovascular or neurological disorders, acute pain, continuous pain or medication for attention deficit hyperactivity disorder, pregnancy and metal implants.

All trauma-exposed subjects fulfilled the trauma criteria of the revised fourth edition of the Diagnostic and Statistical Manual of Mental Disorders (DSM-IV-TR; American Psychiatric Association, 2000) assessed with the German version of the Structured Clinical Interview (SCID-I; Wittchen et al., 1997). Based on the SCID-I and the results of the Clinician-Administered PTSD Scale (CAPS; Blake et al., 1995; Schnyder & Moergeli, 2002), participants were assigned to the respective groups. Healthy control subjects had never experienced any traumatic event in their lives and had never met any criterion for a DSM-IV-TR disorder. All participants received a reimbursement for participation (10€/h), travel and potential costs for accommodation. Patients suffering from PTSD were offered treatment in one of the outpatient clinics of the CIMH.

*Stimuli and experimental procedure*

The virtual contexts were presented on a Dell laptop (Dell Precision M4600; Round Rock, Texas, USA) with a HMD (Trivisio Scout, Kaiserslautern, Rheinland-Pfalz, Germany) outside the scanner and MRI suitable goggles (VisuaStimDigital, Northridge, California, USA) inside the scanner using the same laptop in both cases with a resolution of 800x600 pixels. Ratings were performed on the laptop keyboard during habituation and on a four button optical response pad (Current Designs, Philadelphia, Pennsylvania, USA) during acquisition and extinction.

Each of the four main experimental conditions (context and cue acquisition, context and cue extinction) consisted of 8 room entries per condition following a block design (e.g. context acquisition: 4 x ctx_unpred – 4 x ctx_safe - 4 x cue_pred – 4 x cue_safe). The order of appearance of rooms within each block and of the CSs were counterbalanced using an original (context acquisition: 2 x ctx_unpred-ctx_safe; cue acquisition: 2 x cue_pred-cue_safe) and parallel (context acquisition: 2 x ctx_safe- ctx_unpred; cue acquisition: 2 x cue_safe-cue_pred) version of the experiment. During a pilot study participants had difficulties acquiring the context conditioning with a preceding cue conditioning. We therefore decided to keep the order in which the conditions appeared on each day steady with context acquisition/ extinction appearing before cue acquisition/ extinction (see *Figure 1*).

*Habituation.* During habituation (HAB) participants were passively walked through all four rooms twice and were instructed to pay attention to the interior design of each context. At the end participants were asked “How many different architects designed the rooms?” to guarantee that participants payed attention and could differentiate between the contexts. Furthermore, participants saw each CS separately for 4s with an inter-trial-interval (ITI) of 2s in front of a grey background and included within each context. Participants then rated the arousal, valence and contingency of each context, CS and context with CS on a seven point scale using self-assessment manikins (SAM).. In addition, participants' pain thresholds and the intensity of the painful stimulation were determined (see *Suppl. Methods*), followed by a habituation of the US.

*Acquisition* (see *Figure 1*). During context acquisition, participants were walked through two different contexts, one so called unpredictable (ctx_unpred) and one safe (ctx_safe) context. In the *ctx_unpred* condition, a US was presented at different points in time appearing in the middle of long ITIs between the CTX/CS+/CS- stimuli of 7.5-9 seconds. The CS+ and CS- were presented one to two times per room for four seconds each with additional CTX triggers in each condition. None of the stimuli predicted the appearance of the US in the *ctx_unpred* condition. The *ctx_safe* condition was identical to the *ctx_unpred* except that there was no US presented. At the end of both acquisition phases, participants rated each stimulus (CTX/CS+/CS-) and a combination of them (e.g. CS+ in *ctx_safe*) on the SAM (Bradley & Lang, 1994) for  arousal and valence and the probability of a painful stimulus on a visual analogue scale. During cue acquisition, participants were presented with another two contexts, the so-called predictable (*cue_pred*) and safe (*cue_safe*) context. In the *cue_pred* condition, the 4s presentation of one of the two colored squares (blue/red) was followed by the presentation of the US, which started 0.5 seconds before the end of the CS+. During the *cue_safe* condition, participants received no painful stimulus.

*Extinction.* The extinction phase consisted of the same four conditions, as the acquisition phase (*ctx_unpred_ext*; *ctx_safe_ext*; *cue_pred_ext*; *cue_safe_ext*) except that participants received no painful stimulus in either condition.

*Skin conductance*

The sampling rate was 5000 Hz, filters were DC and 250 Hz, which we downsampled to 10 Hz using Brain Vision Analyzer 2.0 (Brainproducts, Munich, Germany). After manual artefact correction using Ledalab V3.4.9 (http://www.ledalab.de) in Matlab R2016a (The MathWorks Inc., Natick, MA, USA), smoothing with a Gaussian window width of 40 samples, a 6-fold optimization was applied to perform a continuous decomposition analysis. A response window of 1-7.5s after stimulus onset was chosen (Lonsdorf et al., 2017) with a minimum threshold criterion of 0.01 µS. The data was normalized using a logarithmic ($y = \log(x + 1)$) transformation. The electric stimulus was delivered through a cupric (copper) electrode attached to the participants' right hand by an electrical stimulus generator (Digitimer, DS7A, Welwyn Garden City, UK). Increasingly painful stimuli (50 ms bursts, 12 Hz) were administered to participants to obtain the pain threshold and tolerance. The pain intensity and unpleasantness was rated by participants on a Likert scale ranging from 0 (not at all painful/ not at all unpleasant) to 10 (extremely painful/ extremely unpleasant). Participants were asked to rate when a) they felt the electrical stimulus at all, b) when the pain intensity reached seven out of ten points for them and c) when the pain intensity reached an unbearable level (nine out of ten). This procedure was repeated three times in order to obtain a value of 80% of the pain tolerance. To achieve this, the values of the last two runs were entered into the following formula:

$$[(Mean_{pain\_tolerance} - Mean_{pain\_threshold}) * 0.8] + pain\_threshold$$

In case, participants did not rate the pain as aversive after the habituation phase, we increased the intensity by 0.4 mA.

##### *Clinical and neuropsychological assessments and self-reports*

Handedness. The Edinburg Handedness Inventory (EHI; Oldfield, 1971) is a self-report questionnaire in which participant report with which hand they perform a series of sixteen tasks (e.g. writing, holding a spoon). Participants are requested to put a “+” in the column (left hand or right hand) with which they perform the task. If both hands are used for the completion of the task, participants mark this with a “+” in both columns and if exclusively one hand is used, participants mark this with “++” in one of the columns. The “+” are counted and a sum score is built to highlight the dominant hand.

Color-blindness. Participants completed the Ishihara color-blindness test (Ishihara, 1987) which consists of 19 colored dotted items, each depicting a letter, a number or a combination of both. The test separately assesses red-green (15 items) and blue-yellow (three items) color blindness.

Intelligence Testing. The Intelligence score was estimated with a subtest of the Cattell Culture Fair Intelligence Test (CFT, Weiß, 1998) and the „Kurztest für allgemeine Basisgrößen der Informationsverarbeitung“ [Short Test for General Factors of Information Processing] (KAI; Lehrl et al., 1991). In the CFT, participants completed four tests with increasing difficulty. Each test consisted of eight to fourteen questions, in which participants were asked to recognize a pattern/rule within a sequence of figures and apply this rule to either complete the row or figure out “the odd one in the row”. The number of correct responses of all four tests is summed up and the IQ is taken from a table based on a validation sample. In the KAI, participants had to remember a sequence of numbers and digits, starting from three up to the maximum of nine in a row. The test ended when participants could not recall the sequence correctly after the second time.

Posttraumatic Stress Disorder. To assess symptom severity, we used the German version of the Clinician-Administered Posttraumatic Stress Disorder Scale (CAPS; Blake et al., 1995; Schnyder & Moergeli, 2002). The CAPS combined score is calculated by summing the frequency and severity (or intensity) score, measured on two 5-point scales ranging from zero (“never”/ “none) to four (“most or all of the time”/ “extreme”). The CAPS combined score can range from 0 to 100, with either subscore ranging from 0 to 50.

Childhood Trauma Experience. The Childhood Trauma Questionnaire (CTQ; Bernstein et al., 1994) is a 40 item self-report instrument assessing the severity of traumatic childhood experiences, such as emotional abuse and neglect, physical abuse and neglect as well as sexual abuse. The first 34 items ask how often each event occurred during the participant’s upbringing and each item is rated on a 5-point Likert scale ranging from 1 (“never at all”) to 5 (“very often”). For the purpose of this study, we only report the overall sum score, which is calculated by the sum of the five subscales. The overall score can range from 25 to 125. In the last six items, participants are asked to select the age or period in which the neglect or abuse occurred ranging from one to twenty years of age.

Time since trauma and the type of the index event were assessed with the interview on the severity of the trauma [Interview zur Traumaschwere]. The type of traumatic events are hereby subdivided into seven voluntarily (e.g. imprisonment, rape) or five involuntarily (e.g. natural disaster, accident) caused events.

Comorbidities. The German long version of the Center for Epidemiological Studies Depression Scale (ADS; Hautzinger & Bailer, 1993) was applied to assess possible comorbid impairment due to depressive symptoms within the last week. The ADS is a self-report questionnaire with 20 items measured on a 4-point scale ranging from zero (“rarely or not at all [less than one day]”) to three (“most often, all of the time [on five to seven days]”) with a sum score ranging from 0 to 60. Trait anxiety was assessed with the German version of the trait-version of the State-Trait-Anxiety-Inventory (STAI-T; Laux, 1981). The self-report questionnaire comprises of 20 questions, measured on a 4-point Likert scale ranging from one ("not at all") to four ("very much") with higher scores being associated with higher levels of trait anxiety and sum scores ranging from 20 to 80.

Personality Traits. The 60 item version of the Neuroticism-Extraversion-Openness to experience Five-Factor Inventory (NEO-FFI; Costa & McCrae, 2008; Ostendorf & Angleitner, 2004) was used to assess personality traits. Participants can rate each item on a 5-point Likert scale ranging from "strong rejection" to "strong approval". Five trait-dimensions of personality are depicted, namely neuroticism, extraversion, openness to experience, agreeableness and conscientiousness. A sum score is built for each of the five dimensions from twelve items each.

Neuropsychological assessments. Spatial learning and memory were tested with the Cambridge Neuropsychological Test Automated Battery (CANTAB® [Cognitive assessment software]. Cambridge Cognition (2019). All rights reserved. www.cantab.com). First, the Pattern Recognition Test (PRM) is a 2-choice forced discrimination paradigm assessing visual pattern recognition memory. In an initial learning phase, participants are presented with a series of complex visual patterns, one at a time. In a recognition phase, either directly after the testing phase or after a few minutes (delayed), participants have to choose between a novel pattern and a pattern which they have already seen. The outcome variables are the reaction time of a participant's response (mean correct latency) and the accuracy of the responses (percent correct). Second, the Spatial Span (SSP) was assessed with a visuospatial working memory capacity paradigm. Here, participants have to first learn a sequence of two to nine squares that are arranged on the screen and are highlighted in color. They then have to select the squares in the correct order by clicking on the respective squares. When the sequence is correct, participants are presented with an additional square in the next sequence. The outcome variables are the longest sequence successfully recalled (span length), the number of errors (total errors) and the reaction time to the first and last response (speed of response). Third, the Spatial Recognition Memory (SRM) is also a 2-choice forced discrimination paradigm assessing visual-spatial recognition memory. In a learning phase, participants are presented with a sequence of white squares appearing at five different locations on the screen. In the recognition phase, participants are presented with pairs of white squares with one square being in a novel location and one square in a previously shown location. The outcome measures include, similarly to the PRM, the reaction time and accuracy of the responses. Finally, the Paired Associates Learning (PAL) assesses visual memory by showing one to six patterns in a range of white boxes on the screen. Participants have to remember the patterns and location where it appeared. As outcome measures, a memory score is calculated, the mean number of trials to success as well as the total number of trials and errors are measured.

##### *Manipulation check*

Emotional state. Positive and negative affect were measured before and after the acquisition phase on day one and before and after the extinction phase on day two with the Positive And Negative Affective Schedule (PANAS; Watson et al., 1988) and a six item Visual Analogue Scale (VAS). The PANAS has 20 items with ten items each concerning positive and negative affect. Responses are given on a 5-point forced choice scale ranging from 1 (“not at all”) to 5 (“extremely”). The sum scores for each subscale can vary between 10-50 points, with higher scores indicating higher positive/negative affect. The responses on the VAS ranged from 0 (“applies not at all”) to 10 (“applies completely”) with six items describing the current mood: 1) “high mood”, 2) “irritated”, 3) “balanced”, 4) “gloomy mood”, 5) “sluggish”, 6) “activated”.

Debriefing. A set of seven questions were asked at the end of habituation and acquisition on day one of the experiment. The questions were the following: 1) "How many different architects designed the rooms?", 2) "How quickly did you manage to distinguish the rooms from each other?" with possible responses being "during context acquisition/ during cue acquisition/ not at all", 3) "Did you find the instructions understandable?" with responses ranging from 1 ("difficult") to 10 ("easy"), 4) "Did you find the ratings understandable?" with responses ranging from 1 ("difficult") to 10 ("easy"), 5) "How well did you get along with the keyboard?" with responses ranging from 1 ("very badly") to 10 ("very good"), 6) "How exhausting did you find the experiment?" from 1 ("very exhausting") to 10 ("not exhausting at all"), 7) "How attentive were you during the experiment?" from 1 ("not at all") to 10 ("very").

#### *MRI data acquisition*

Blood-oxygenation-level-dependent (BOLD) contrasts of whole-brain functional images were acquired using a T2*-weighted Gradient-Echo-Planar Imaging (EPI) sequence (protocol parameters: TR = 2700 ms; TE = 27 ms; matrix size = 96 x 96; field of view = 220 x 220 $mm^2$; flip angle = 90°; GRAPPA PAT 2; sequence length: 19:02min). Each of the 420 volumes per condition consisted of 40 axial slices (slice thickness = 2.3 mm; gap = 0.7 mm; voxel size = 2.3 $mm^3$) measured in interleaved, descending slice order and positioned along a tilted line to the anterior-posterior commissure (AC-PC orientation). An automated high-order shimming technique was used to maximize magnetic field homogeneity.

#### *Statistical analysis*

Behavioral, SCR. For the analyses of the self-report ratings (arousal, valence, contingency) and the SCR we performed two separate analyses each. In case of the self-report ratings, we performed a 3 (Groups) x 3 (Phase: HAB, ACQ, EXT) repeated measures ANOVA (rmANOVA) for each of the four contexts. For the difference scores (CS+-CS-) of the ratings, we performed two separate 3 (Groups) x 2 (condition: context or cue) rmANOVAs, one for the acquisition and one for the extinction phase. In case of the SCRs, we performed 3 (Groups) x 2 (contexts: e.g. ctx_unpred and ctx_safe) rmANOVAs, one for each phase (ACQ or EXT) and condition (context or cue). The difference scores (CS+-CS-) of the SCRs were calculated in a similar fashion with four separate 3 (Groups) x 2 (contexts: e.g. ctx_unpred and ctx_safe) rmANOVAs.

fMRI. A response window of 1-7s after stimulus onset for all parameters (ctx, cs+, cs-) was chosen for BOLD responses. We then extracted beta values from the first level from a priori defined ROIs, namely the hippocampi, the amygdalae and the vmPFC. The masks were taken from the Wake Forest University Pick Atlas 3.0.5b (Maldjian et al., 2003) choosing bilaterally the hippocampi and amydalae from the Automated Anatomical Labeling Atlas (AAL; Tzourio-Mazoyer et al., 2002). For the vmPFC, we chose Brodmann areas (BA) 11, 12 and 25 (Wicking et al., 2016). Beta values were extracted directly from SPM12 with customized MATLAB scripts.

On the first level, we set up four different general linear models (GLM), one for each phase (context acquisition, cue acquisition, context extinction, cue extinction) including the following six experimental predictors in each model: 1) conditioned fear context [CTX+], 2) conditioned safety context [CTX-], 3) conditioned fear cue in fear context [CS+ in CTX+], 4) conditioned fear cue in safety context [CS+ in CTX-], 5) conditioned safety cue in fear context [CS- in CTX+], 6) conditioned safety cue in safety context [CS- in CTX-]. As an example, for the context conditioning phase this resulted in the following six predictors: ctx_unpred, ctx_safe, cs+ (in ctx_unpred), cs+ (in ctx_safe), cs- (in ctx_unpred), cs- (in ctx_safe). In addition, each model contained six parameters describing the rigid body transformation to account for head motion (in mm: x-, y-, z-direction; in degrees: pitch-, roll-, yaw-direction).

The fMRI data was analyzed using Statistical Parametric Mapping (SPM12; Wellcome Department of Imaging Neuroscience, London, UK) implemented in MATLAB R2016a (The MathWorks Inc., Natick, MA, USA). Before preprocessing, the first five volumes of each scanning session were discarded to allow for $T_1$ equilibration effects. Participants were excluded in case their motion parameter estimates exceeded 2.3 mm in x-, y-, or z-direction and a maximum of 1° of any angular motion throughout the course of the scan. Preprocessing included realignment, normalization to the standard space of the Montreal Neurological Institute (MNI; SPM12 template), slice time correction to reference slice one, coregistration of structural and functional volumes and smoothing of each functional volume with a 8.0 x 8.0 x 8.0 $mm^3$ Gaussian kernel. We extracted beta values and compared them for the following, a priori, defined ROIs: hippocampi, amygdalae, vmPFCs. In addition, we performed exploratory whole brain contrasts.

### Supplemental results

#### *Sample characteristics*

Demographic Information. All detailed information can be found in *Table 1.* The sample did not significantly differ in the distribution of gender ($X^2$(2, 63) = 0.29, *p* = .87) with approximately 50% females in each group, nor in age (*F*(2, 60) = 1.09, *p* = .34) with participants' age ranging from 20 to 62 years across groups. The groups did significantly differ in the level of education ($X^2$(2, 62) = 9.67, *p* = .008) whereby the distribution of the level of education of patients with PTSD ($N_{\leq 12}$=12/ $N_{>12}$=7) was significantly lower from the two control groups ($p_{bonf.cor.}$ = .015) as assessed by a chi-square post-hoc test. Finally, there were no significant differences between the groups in the distribution of handedness ($X^2$(4, 62) = 1.03, *p* = .91), or the intelligence quotients as assessed with the KAI (*F*(2, 56) = 2.92, *p* = .06; range 82 to 142) and the CFT (*F*(2, 57) = 0.94, *p* = .40; range 69 to 140; see *Table 1* for details).

Trauma severity. All detailed information can be found in *Table 1.* Time since trauma did not significantly differ between patients with PTSD and TC subjects (*T*(24.7) = 1.75, *p* = .09; 95% CI -.96 to 11.85), ranging from 1 to 30 years. The groups did also not significantly differ across the two types of traumatic events ($X^2$(1, 41) = 0.61, *p* = .44; see *Table 1* for details).

Trauma diagnostics. All detailed information can be found in *Table 1.* Patients with PTSD showed a significantly higher overall CAPS score than TC subjects (*T*(36.9) = 9.02, *p* < .001; 95% CI -60.38 to 38.22) as well as significantly higher CAPS severity (*T*(37.9) = 7.20, *p* < .001; 95% CI -29.34 to -16.46) and CAPS frequency (*T*(35.1) = 9.56, *p* < .001; 95% CI -31.58 to -20.52) scores. There was a significant difference in the CAPS score between the experimental groups (*F*(2, 59) = 3.27, *p* = .045), with patients with PTSD showing significantly higher scores than TC subjects ($M_{Difference}$ = 11.2; 95% CI 0.7 to 21.8, *p* = .035; *Hedges'g* = 0.77; see *Table 1* for details).

Comorbidities. All detailed information can be found in *Table 1.* Patients with PTSD showed significantly higher numbers of comorbidities than both control groups, both on axis I ($X^2(2, 63) = 21.05, p < .001$) and on axis II disorders ($X^2(2, 63) = 14.26, p < .001$). Comorbidities on Axis I disorders were comprised of current major depressive disorder (MDD; $N_{PTSD} = 10$), previous MDD ($N_{PTSD} = 8$; $N_{TC} = 2$), general anxiety disorder (GAD; $N_{PTSD} = 2$), panic disorder ($N_{PTSD} = 4$; $N_{TC} = 2$) substance dependence ($N_{PTSD} = 2$; $N_{TC} = 1$) and alcohol abuse ($N_{PTSD} = 1$; $N_{TC} = 2$), previous manic episode ($N_{PTSD} = 2$), current dysthymia ($N_{PTSD} = 1$) and bulimia ($N_{PTSD} = 1$). Comorbidities on Axis II disorders for patients with PTSD comprised of avoidant personality disorder ($N_{PTSD} = 4$), obsessive-compulsive personality disorder ($N_{PTSD} = 1$) and depressive personality disorder ($N_{PTSD} = 1$; see *Table 1* for details). The groups differed significantly in their depression score ($F(2, 59) = 23.58, p < .001$; range 0 to 42) with post-hoc tests revealing significantly higher depression in patients with PTSD compared to TC ($M_{Difference} = 11.8$; 95% CI 4.8 to 18.8, $p_{Tukey\ HSD} < .001$; *Hedges'g* = 1.08), between patients with PTSD and HC ($M_{Difference} = 19.7$; 95% CI 12.8 to 26.6, $p_{Tukey\ HSD} < .001$; *Hedges'g* = 2.51) as well as between TC and HC ($M_{Difference} = 7.9$; 95% CI 1.2 to 14.6, $p_{Tukey\ HSD} = .018$; *Hedges'g* = 0.94; see *Table 1* for details). The groups also differed significantly in their STAI-T score ($F(2, 59) = 20.28, p < .001$; range 23 to 70) with post-hoc tests revealing a significantly higher STAI-T score between patients with PTSD and TC($M_{Difference} = 12.1$; 95% CI 4.0 to 20.2, $p_{Tukey\ HSD} = .002$; *Hedges'g* = 0.99), between patients with PTSD and HC ($M_{Difference} = 21.2$; 95% CI 13.2 to 29.2, $p_{Tukey\ HSD} < .001$; *Hedges'g* = 2.17) and between TC and HC ($M_{Difference} = 9.1$; 95% CI 1.3 to 16.9, $p_{Tukey\ HSD} = .019$; *Hedges'g* = 0.89; see *Table 1* for details). Finally, the experimental groups did not significantly differ in the distribution of intake of any prescribed medication ($X^2(2, 63) = 2.18, p = .34$), with three subjects reporting the intake of low doses of psychopharmacological medication ($N_{PTSD} = 1$; $N_{TC} = 2$; longterm usage of Tetrahydrocannabinol, Pregabalin, Quetiapin), five subjects reporting the intake of nonpsychopharmacological medication ($N_{PTSD}$ = 2; $N_{TC}$ = 2; $N_{HC}$ = 1; contraceptive pill, L-Thyroxine, Mesalazine, Prednisolone) and 55 subjects reporting no intake of any medication; see *Table 1* for details).

Personality traits. All detailed information can be found in *Suppl. Table 1.* The experimental groups did not significantly differ on extraversion ($F(2, 55) = 3.03, p = .056$; range 6 to 43), openness to experience ($F(2, 57) = 0.77, p = .47$; range 10 to 45) and conscientiousness ($F(2, 58) = 2.23, p = .12$; range 17 to 45). They did, however, significantly differ on neuroticism ($F(2, 56) = 12.87, p < .001$; range 2 to 37), with post-hoc tests revealing a significantly higher neuroticism score for patients with PTSD versus TC ($M_{Difference}$ = 7.6; 95% CI 1.1 to 14.1, $p_{Tukey\ HSD} = .019$; *Hedges'g* = 0.89), for patients with PTSD compared to HC ($M_{Difference}$ = 13.9; 95% CI 7.3 to 20.4, $p_{Tukey\ HSD} < .001$; *Hedges'g* = 1.77) and a marginally significantly higher score for TC versus HC ($M_{Difference}$ = 6.3; 95% CI 0.0 to 12.6, $p_{Tukey\ HSD} = .051$; *Hedges'g* = 0.77). The groups did also significantly differ on agreeableness ($F(2, 56) = 4.98, p = .010$; range 16 to 46), with post-hoc tests revealing significantly lower agreeableness scores for patients with PTSD versus HC ($M_{Difference}$ = -6.1; 95% CI -11.2 to -1.0, $p_{Tukey\ HSD} = .014$; *Hedges'g* = 0.92) and for TCs versus HCs ($M_{Difference}$ = -5.0; 95% CI -9.8 to -0.1, $p_{Tukey\ HSD} < .045$; *Hedges'g* = 0.79), with no significant difference between patients with PTSD and TC ($M_{Difference}$ = -1.2; 95% CI -6.4 to 4.1, $p_{Tukey\ HSD} = .86$; *Hedges'g* = 0.16; see *Suppl. Table 1* for details).

Neuropsychological Assessment. There was no significant difference between patients with PTSD, TC and HC subjects in any of the scores of the PRM, PRM delayed, SSP, SRM or PAL (see *Suppl. Table 1* for details).

Debriefing. All detailed information can be found in *Suppl. Table 2*. The groups did not significantly differ on any of the debriefing questions. After habituation, the groups reported a similar number of architects designing the rooms ($F(2, 49) = 2.62, p = .08$; range 2 to 9). After acquisition, the groups did not significantly differ on when they could distinguish the contexts ($X^2$(4, 61) = 6.07, $p$ = .19) with the majority of participants (67%) being able to distinguish the rooms from each other during context acquisition. In addition, participants across all groups found the instructions ($F$(2, 59) = 0.53, $p$ = .39; range 5 to 10) and ratings ($F$(2, 60) = 2.65, $p$ = .11; range 3 to 10) understandable, could handle the keyboard ($F$(2, 60) = 1.58, $p$ = .21; range 1 to 10) and were similarly exhausted after ($F$(2, 58) = 2.26, $p$ = .14; range 1 to 10) and attentive during ($F$(2, 59) = 0.07, $p$ = .80; range 1 to 10) the experiment (see *Suppl. Table 2* for details).

#### *Self-reports*

Ratings of the unconditioned stimulus. All detailed information can be found in *Suppl. Table 3a*). The experimental groups did not significantly differ in the ratings of the intensity of the US ($F$(2, 58) = 0.54, $p$ = .59) at the end of habituation. For the pain intensity rating, there was a significant main effect of phase ($F_{phase}$(2, 108) = 31.27, $p$ < .001) and a significant interaction of phase x group ($F_{group\ x\ phase}$(4, 108) = 3.05, $p$ = .02). The pain intensity ratings of the US were higher during habituation than during context and cue conditioning across all three groups. However, the pain intensity ratings for the US were higher for TC subjects than patients with PTSD and HC subjects during context and cue conditioning. For the valence ratings of the US, we found a significant main effect of phase ($F_{phase}$(2, 108) = 18.46, $p_{GG}$ < .001) and a significant interaction of group x phase ($F_{group\ x\ phase}$(4, 108) = 3.06, $p_{GG}$ = .031). Similar to the pain intensity ratings, the valence ratings of the US were higher for the habituation than for cue and context conditioning across all three groups. However, the valence ratings of the US stayed higher for TC subjects than patients with PTSD and HC subjects during cue and context conditioning (*Suppl. Table 3a*).

Ratings across contexts. All detailed information can be found in *Suppl. Figure 2a, Suppl. Table 3b-d*. We found a significant main effect of phase for arousal ratings in the *ctx_unpred* ($F_{phase}(2, 82) = 4.33$, $p = .022$), ctx_safe ($F_{phase}(1, 46) = 6.45$, $p = .015$), *cue_pred* ($F_{phase}(2, 82) = 6.03$, $p = .004$) and *cue_safe* ($F_{phase}(1, 46) = 9.55$, $p = .003$) condition. The arousal ratings were highest after acquisition across all the groups. There was no significant main effect of group and no significant interaction of group x phase in the arousal ratings (*Suppl. Table 3b*). In the valence rating for each context, we only found a significant main effect of phase for the *ctx_unpred* ($F_{phase}(2, 82) = 8.90$, $p < .001$) and a significant interaction of group x phase in *ctx_safe* ($F_{group\ x\ phase}(2, 46) = 3.69$, $p = .033$; *Suppl. Table 3c*). The valence ratings were highest after acquisition across all the groups for *ctx_unpred*. For ctx_safe, the valence ratings were higher during acquisition than extinction, while for TC subjects it was the opposite, hence the significant interaction. For the contingency ratings, we observed significant main effects of phase across all four contexts, namely *ctx_unpred* ($F_{phase}(2, 82) = 10.56$, $p < .001$), *ctx_safe* ($F_{phase}(1, 46) = 13.36$, $p < .001$), *cue_pred* ($F_{phase}(2, 82) = 5.37$, $p = .007$) and *cue_safe* ($F_{phase}(1, 46) = 16.57$, $p < .001$). The contingency ratings were highest after acquisition across all the groups and contexts.

Differences in ratings between CS+ - CS-. All detailed information for the difference scores of CS+ - CS- can be found in *Suppl. Figure 2b* and *Suppl. Table 3e*. A significant main effect of phase was found for the difference ratings during acquisition for arousal ($F_{phase}(1, 56) = 39.13$, $p < .001$), valence ($F_{phase}(1, 56) = 21.54$, $p < .001$) and contingency ($F_{phase}(1, 56) = 42.89$, $p < .001$). All three groups seemed to be able to recognize the CS+ as danger signal during cue conditioning in the predictable context.

##### *Clinical Correlations*

ACQ. Pearson correlations were performed between the CAPS sumscore and the Left hippocampus (r(33)=-.17, p=.33) and right hippocampus (r(33)=.08, p=.64) during acquisition of context unpredictable.

EXT. Pearson correlations were also performed between the sumscore of the CAPS and the left vmPFC (r(33)=.30, p=.08) and right vmPFC (r(33)=.27, p=.12) during extinction of context unpredictable as well as the sumscore of the CAPS and the left vmPFC (r(33)=-.20, p=.25) or right vmPFC (r(33)=-.25, p=.16) during extinction of context predictable.

None of the correlation surpassed the significance level applying multiple comparison Bonferroni correction (alpha/6=.008).

| | | Groups | | | | | | | | | | | | Analyses | | | | | | | | | |
|---|---|---|---|---|---|---|---|---|---|---|---|---|---|---|---|---|---|---|---|---|---|---|---|
| | | **PTSD** [N=20] | | | | **TC** [N=21] | | | | **HC** [N=22] | | | | | | | | | | | | | |
| | | *M* | *SD* | *n* | *(%)* | *M* | *SD* | *n* | *(%)* | *M* | *SD* | *n* | *(%)* | $X^2$ | *F* | *T* | *Df* | *p* | *Cont.* | *Diff,* | *CI [-95%; +95%]* | $p_{Tukey HSD}$ | *Hedges' g* |
| NEO-FFI - Neuroticism | | 26.6 | 9.2 | 18 | | 19.0 | 9.4 | 21 | | 12.8 | 6.2 | 20 | | | 12.87 | | 2 | <.001 | T-H | 6.3 | -0.0; 12.6 | .051 | 0.77 |
| | | | | | | | | | | | | | | | | | | | P-H | 13.9 | 7.3; 20.4 | <.001 | 1.77 |
| | | | | | | | | | | | | | | | | | | | P-T | 7.6 | 1.1; 14.1 | .019 | 0.89 |
| NEO-FFI - Extraversion | | 22.9 | 8.2 | 16 | | 26.7 | 7.2 | 21 | | 29.1 | 7.4 | 21 | | | 3.03 | | 2 | .056 | | | | | |
| NEO-FFI - Openness to experience | | 27.9 | 9.3 | 17 | | 31.2 | 7.5 | 21 | | 29.5 | 7.9 | 22 | | | 0.77 | | 2 | .47 | | | | | |
| NEO-FFI - Agreeableness | | 31.4 | 7.1 | 17 | | 32.5 | 6.4 | 20 | | 37.5 | 6.3 | 22 | | | 4.98 | | 2 | .010 | T-H | -5.0 | -9.8; -0.1 | .045 | 0.79 |
| | | | | | | | | | | | | | | | | | | | P-H | -6.1 | -11.2;-1.0 | .014 | 0.92 |
| | | | | | | | | | | | | | | | | | | | P-T | -1.2 | -6.4; 4.1 | .86 | 0.16 |
| NEO-FFI - Conscientiousness | | 30.6 | 8.0 | 18 | | 32.6 | 5.3 | 21 | | 34.8 | 5.8 | 22 | | | 2.23 | | 2 | .12 | | | | | |
| **Neuropsychological Assessments** | | | | | | | | | | | | | | | | | | | | | | | |
| PRM | Mean correct latency | 2.45 | 0.74 | 18 | | 2.11 | 0.59 | 21 | | 2.19 | 0.53 | 21 | | | 1.60 | | 2 | .21 | | | | | |
| | Percent correct | 90.3 | 10.4 | 18 | | 94.8 | 4.15 | 21 | | 94.0 | 9.18 | 21 | | 12.64 | | | 8 | .13 | | | | | |
| PRM delayed | Mean correct latency | 2.13 | 0.49 | 18 | | 2.02 | 0.55 | 21 | | 2.24 | 1.27 | 21 | | | 0.36 | | 2 | .70 | | | | | |
| | Percent correct | 75.5 | 16.0 | 18 | | 85.2 | 13.1 | 21 | | 82.9 | 14.8 | 21 | | 13.66 | | | 16 | .62 | | | | | |
| SSP | Span length | 5.61 | 1.85 | 18 | | 6.57 | 1.21 | 21 | | 6.63 | 1.34 | 19 | | | 2.79 | | 2 | .07 | | | | | |
| | Total errors | 12.8 | 5.87 | 18 | | 15.1 | 7.11 | 21 | | 13.8 | 5.64 | 19 | | | 0.67 | | 2 | .52 | | | | | |
| | Mean time to first response | 2.42 | 0.66 | 18 | | 2.73 | 0.40 | 21 | | 2.90 | 0.69 | 19 | | | 3.07 | | 2 | .055 | | | | | |
| | Mean time to last response | 3.14 | 1.03 | 18 | | 3.50 | 0.63 | 21 | | 3.58 | 0.73 | 19 | | | 1.57 | | 2 | .22 | | | | | |
| | Total usage errors | 2.72 | 2.11 | 18 | | 2.24 | 1.34 | 21 | | 2.53 | 1.95 | 19 | | | 0.36 | | 2 | 0.70 | | | | | |
| SRM | Mean correct latency | 2.00 | 0.44 | 18 | | 2.09 | 0.58 | 21 | | 2.06 | 0.51 | 21 | | | 0.17 | | 2 | .84 | | | | | |
| | Percent correct | 78.3 | 10.6 | 18 | | 79.3 | 8.70 | 21 | | 76.7 | 9.79 | 21 | | 17.09 | | | 18 | .52 | | | | | |
| PAL | First trial memory score | 19.0 | 2.68 | 18 | | 20.6 | 3.75 | 21 | | 20.6 | 5.31 | 21 | | | 0.95 | | 2 | .39 | | | | | |
| | Mean trials to success | 1.62 | 0.44 | 18 | | 1.46 | 0.32 | 21 | | 1.48 | 0.62 | 21 | | | 0.62 | | 2 | .54 | | | | | |
| | Total error (adjusted) | 15.6 | 12.6 | 18 | | 9.76 | 8.50 | 21 | | 12.0 | 16.3 | 21 | | | 1.01 | | 2 | .37 | | | | | |
| | Total trials | 12.8 | 3.26 | 18 | | 11.7 | 2.59 | 21 | | 11.6 | 4.25 | 21 | | | 0.68 | | 2 | .51 | | | | | |

**Suppl. Table 1.** Assessment of personality traits and Neuropsychological assessments.

**[Abbreviations:** Cont. – Contrast; H – Healthy control subjects; NEO-FFI – Neuroticism-Extraversion-Openness to experience Five-Factor Inventory; PAL – Paired Associates Learning; PRM – Pattern Recognition Memory; P – patients with PTSD; SRM – Spatial Recognition Memory; SSP – Spatial Span; T – Trauma control subjects]

| Debriefing | | PTSD | | | | TC | | | | HC | | | | Analysis | | | |
|---|---|---|---|---|---|---|---|---|---|---|---|---|---|---|---|---|---|
| | | *M* | *SD* | *n* | *%* | *M* | *SD* | *n* | *%* | *M* | *SD* | *n* | *%* | $X^2$ | *F* | *Df* | *p* |
| 1. How many different architects designed the rooms? | | 6.04 | 2.37 | 14 | | 6.69 | 1.76 | 18 | | 5.25 | 1.77 | 20 | | | 2.62 | 2 | .08 |
| 2. How quickly did you manage to distinguish the rooms from each other? | During Context ACQ | | | 10 | 52.6 | | | 14 | 70.0 | | | 17 | 77.3 | 6.07 | | 4 | .19 |
| | During Cue ACQ | | | 7 | 36.9 | | | 6 | 30.0 | | | 5 | 22.7 | | | | |
| | Not at all | | | 2 | 10.5 | | | 0 | 0.0 | | | 0 | 0.0 | | | | |
| 3. Did you find the instructions understandable? [1 “difficult”; 10 “easy”] | | 9.00 | 1.34 | 20 | | 9.7 | 0.80 | 20 | | 9.24 | 1.22 | 21 | | | 0.53 | 1 | .39 |
| 4. Did you find the ratings understandable? [1 “difficult”; 10 “easy”] | | 8.15 | 1.87 | 20 | | 8.7 | 1.72 | 20 | | 8.95 | 1.17 | 22 | | | 2.65 | 1 | .11 |
| 5. How well did you get along with the keyboard? [1 “very badly”; 10 “very good”] | | 8.80 | 1.61 | 20 | | 8.35 | 2.32 | 20 | | 9.45 | 1.06 | 22 | | | 1.58 | 1 | .21 |
| 6. How exhausting did you find the experiment? [1 “very exhausting”; 10 “not exhausting at all”] | | 5.00 | 2.81 | 19 | | 6.40 | 2.04 | 20 | | 6.14 | 2.10 | 21 | | | 2.26 | 1 | .14 |
| 7. How attentive were you during the experiment? [1 “not at all”; 10 “very”] | | 7.21 | 2.15 | 19 | | 7.65 | 1.53 | 20 | | 7.36 | 1.33 | 22 | | | 0.07 | 1 | .80 |

**Supplementary Table 2.** Results of debriefing questionnaire asked at the end of the habituation (Question 1) and at the end of acquisition (Question 2-7).

[**Abbreviations:** HC – Healthy control subjects without trauma experience; PTSD – patients with PTSD; TC – healthy control subjects with trauma experience]

# US

| Groups | n | HAB<br>M (SD) | ACQ Con<br>M (SD) | ACQ Cue<br>M (SD) | Analyses |
|---|---|---|---|---|---|
| **Intensity (in mA)** | | | | | |
| PTSD | [n=19] | 4.68 | - | - | F(2, 58)= 0.54, p=.59 |
| | | (3.39) | | | |
| TC | [n=20] | 4.80 | - | - | |
| | | (2.41) | | | |
| HC | [n=22] | 3.96 | - | - | |
| | | (2.72) | | | |
| **Pain** | | | | | |
| PTSD | [n=17] | 7.29 | 5.47 | 5.94 | Group: F(2, 54)= 2.29, p=.11 |
| | | (0.77) | (1.94) | (1.71) | **Phase: F(2, 108)= 31.27, p<.001*** ** |
| TC | [n=20] | 7.10 | 6.55 | 6.55 | HAB > $ACQ_{Con}$ + $ACQ_{Cue}$ |
| | | (0.45) | (0.76) | (0.76) | **GroupxPhase: F(4, 108)= 3.05, p=.02*** |
| HC | [n=20] | 7.25 | 5.40 | 5.50 | $TC_{ACQ\_con}$ > $PTSD_{ACQ\_con}$ + $HC_{ACQ\_con}$ |
| | | (0.44) | (2.06) | (1.82) | $TC_{ACQ\_cue}$ > $PTSD_{ACQ\_cue}$ + $HC_{ACQ\_cue}$ |
| **Valence** | | | | | |
| PTSD | [n=17] | 7.29 | 5.76 | 6.18 | Group: F(2, 54)= 3.02, p=.057 |
| | | (0.69) | (1.92) | (1.88) | **Phase: F(2, 108)= 18.46, $p_{GG}$<.001*** ** |
| TC | [n=20] | 7.10 | 6.75 | 6.85 | HAB > $ACQ_{Con}$ + $ACQ_{Cue}$ |
| | | (0.45) | (1.02) | (1.18) | **GroupxPhase: F(4, 108)= 3.06, $p_{GG}$=.031*** |
| HC | [n=20] | 7.10 | 5.60 | 5.35 | $TC_{ACQ\_con}$ > $PTSD_{ACQ\_con}$ + $HC_{ACQ\_con}$ |
| | | (0.55) | (1.90) | (2.21) | $TC_{ACQ\_cue}$ > $HC_{ACQ\_cue}$ |

**Suppl. Table 3a.** Intensity (in Milliampere), pain intensity ratings and valence ratings of the US during HAB and ACQ.

[**Abbreviations:** ACQ – Acquisition; Con – Context; EXT – Extinction; HAB – Habituation; HC – Healthy control subjects without trauma experience; mA – Milliampere; PTSD – patients with PTSD; TC – healthy control subjects with trauma experience; US – Unconditioned Stimulus]

#### Arousal

| Groups | n | HAB M (SD) | ACQ M (SD) | EXT M (SD) | Analyses |
|---|---|---|---|---|---|
| **CTX_unpred** | | | | | |
| PTSD | [n=14] | 2.21 | 2.46 | 1.54 | Group: F(2, 41)= 1.40, p=.26 |
| | | (1.73) | (1.68) | (0.91) | **Phase: F(2, 82)= 4.33, $p_{GG}$=.022*** |
| TC | [n=17] | 2.41 | 3.47 | 2.24 | ACQ > HAB + EXT |
| | | (1.24) | (2.22) | (1.44) | GroupxPhase: F(4, 82)= 1.38, p=.25 |
| HC | [n=13] | 2.15 | 2.27 | 2.23 | |
| | | (1.25) | (0.90) | (1.13) | |
| **CTX_safe** | | | | | |
| PTSD | [n=16] | - | 2.31 | 1.69 | Group: F(2, 46)= 0.80, p=.46 |
| | | | (1.52) | (1.40) | **Phase: F(1, 46)= 6.45, p=.015*** |
| TC | [n=18] | - | 2.72 | 2.42 | ACQ > EXT |
| | | | (1.22) | (2.10) | GroupxPhase: F(2, 46)= 0.33, p=.72 |
| HC | [n=15] | - | 2.60 | 1.90 | |
| | | | (1.45) | (1.14) | |
| **CUE_pred** | | | | | |
| PTSD | [n=14] | 2.46 | 2.83 | 2.00 | Group: F(2, 42)= 1.30, p=.28 |
| | | (1.83) | (1.83) | (1.40) | **Phase: F(2, 82)= 6.03, p=.004**** |
| TC | [n=17] | 2.50 | 3.62 | 2.79 | ACQ > HAB + EXT |
| | | (1.35) | (2.33) | (2.27) | GroupxPhase: F(4, 82)= 0.56, p=.69 |
| HC | [n=13] | 1.85 | 2.62 | 2.04 | |
| | | (0.90) | (1.34) | (1.25) | |
| **CUE_safe** | | | | | |
| PTSD | [n=16] | - | 2.28 | 1.78 | Group: F(2, 46)= 0.34, p=.72 |
| | | | (1.48) | (1.03) | **Phase: $F_P$(1, 46)= 9.55, p=.003**** |
| TC | [n=18] | - | 2.56 | 2.08 | ACQ > EXT |
| | | | (1.12) | (1.41) | GroupxPhase: F(2, 46)= 0.12, p=.89 |
| HC | [n=15] | - | 2.63 | 1.97 | |
| | | | (1.25) | (1.34) | |

**Supplementary Table 3b.** Mixed repeated measures ANOVAs (rmANOVA) across arousal ratings for each of the four conditions (ctx_unpred, ctx_safe, cue_pred, cue_safe) and each of the three phases (HAB, ACQ, EXT).

[**Abbreviations:** ACQ – Acquisition; CTX – Context; EXT – Extinction; HAB – Habituation; HC – Healthy control subjects without trauma experience; $p_{GG}$ – Greenhouse-Geisser correction; pred – Predictable; PTSD – patients with PTSD; SCR – Skin conductance response; TC – healthy control subjects with trauma experience; unpred – Unpredictable]

### Valence

| Groups | n | HAB M (SD) | ACQ M (SD) | EXT M (SD) | Analyses |
|---|---|---|---|---|---|
| **CTX_unpred** | | | | | |
| PTSD | [n=14] | 3.54 (1.67) | 3.71 (1.99) | 3.36 (1.79) | Group: $F(2, 41)= 0.30, p=.74$<br>**Phase: $F(2, 82)= 8.90, p_{GG}<.001$***** |
| TC | [n=17] | 3.26 (1.38) | 4.18 (2.08) | 3.18 (1.49) | ACQ > HAB + EXT<br>GroupxPhase: $F(4, 82)= 0.92, p=.45$ |
| HC | [n=13] | 2.81 (1.49) | 3.85 (1.63) | 2.81 (1.63) | |
| **CTX_safe** | | | | | |
| PTSD | [n=16] | - | 3.53 (1.79) | 3.22 (1.81) | Group: $F(2, 46)= 0.86, p=.43$<br>Phase: $F(1, 46)= 2.26, p=.14$ |
| TC | [n=18] | - | 3.33 (1.37) | 3.72 (2.12) | **GroupxPhase: $F(2, 46)= 3.69, p=.033$***<br>$HC_{EXT} > PTSD_{EXT} + TC_{EXT}$ |
| HC | [n=15] | - | 3.40 (1.66) | 2.33 (0.96) | |
| **CUE_pred** | | | | | |
| PTSD | [n=14] | 3.89 (1.91) | 3.63 (1.91) | 4.00 (2.12) | Group: $F(2, 42)= 0.19, p=.83$<br>Phase: $F(2, 82)= 0.19, p=.83$ |
| TC | [n=17] | 3.65 (1.43) | 3.65 (1.94) | 3.88 (2.18) | GroupxPhase: $F(4, 82)= 1.08, p=.37$ |
| HC | [n=13] | 3.37 (1.91) | 3.81 (1.68) | 2.96 (1.89) | |
| **CUE_safe** | | | | | |
| PTSD | [n=16] | - | 3.69 (1.71) | 3.84 (1.94) | Group: $F(2, 46)= 1.83, p=.17$<br>Phase: $F_P(1, 46)= 0.01, p=.94$ |
| TC | [n=18] | - | 3.25 (1.31) | 3.61 (1.92) | GroupxPhase: $F(2, 46)= 1.29, p=.29$ |
| HC | [n=15] | - | 3.00 (1.21) | 2.53 (1.70) | |

**Supplementary Table 3c.** Mixed repeated measures ANOVAs (rmANOVA) across valence ratings for each of the four conditions (ctx_unpred, ctx_safe, cue_pred, cue_safe) and each of the three phases (HAB, ACQ, EXT).

[**Abbreviations:** ACQ – Acquisition; CTX – Context; EXT – Extinction; HAB – Habituation; HC – Healthy control subjects without trauma experience; $p_{GG}$ – Greenhouse-Geisser correction; pred – Predictable; PTSD – patients with PTSD; SCR – Skin conductance response; TC – healthy control subjects with trauma experience; unpred – Unpredictable]

#### Contingency

| Groups | n | HAB M (SD) | ACQ M (SD) | EXT M (SD) | Analyses |
|---|---|---|---|---|---|
| **CTX_unpred** | | | | | |
| PTSD | [n=14] | 2.64 | 3.68 | 2.11 | Group: F(2, 41)= 1.12, p=.34 |
| | | (1.70) | (1.87) | (2.26) | **Phase: F(2, 82)= 10.56, $p_{GG}$<.001*** ** |
| TC | [n=17] | 3.12 | 4.29 | 2.38 | EXT < HAB + ACQ |
| | | (1.60) | (1.98) | (1.75) | GroupxPhase: F(4, 82)= 0.55, p=.70 |
| HC | [n=13] | 3.08 | 3.12 | 1.58 | |
| | | (1.80) | (2.58) | (1.10) | |
| **CTX_safe** | | | | | |
| PTSD | [n=16] | - | 3.22 | 2.06 | Group: F(2, 46)= 0.92, p=.41 |
| | | | (1.91) | (2.15) | **Phase: F(1, 46)= 13.36, p<.001*** ** |
| TC | [n=18] | - | 3.64 | 2.64 | ACQ > EXT |
| | | | (1.75) | (2.17) | GroupxPhase: F(2, 46)= 0.42, p=.66 |
| HC | [n=15] | - | 3.33 | 1.57 | |
| | | | (2.53) | (0.89) | |
| **CUE_pred** | | | | | |
| PTSD | [n=14] | 2.89 | 2.75 | 1.89 | Group: F(2, 42)= 2.65, p=.83 |
| | | (1.96) | (2.03) | (1.71) | **Phase: F(2, 82)= 5.27, p=.007** ** |
| TC | [n=17] | 3.41 | 4.24 | 2.79 | EXT < HAB + ACQ |
| | | (1.65) | (2.22) | (2.28) | GroupxPhase: F(4, 82)= 0.53, p=.72 |
| HC | [n=13] | 2.81 | 2.62 | 1.85 | |
| | | (1.74) | (1.96) | (1.39) | |
| **CUE_safe** | | | | | |
| PTSD | [n=16] | - | 2.78 | 1.56 | Group: F(2, 46)= 0.99, p=.38 |
| | | | (1.83) | (1.14) | **Phase: $F_P$(1, 46)= 16.57, p<.001*** ** |
| TC | [n=18] | - | 2.94 | 2.17 | ACQ > EXT |
| | | | (1.70) | (1.56) | GroupxPhase: F(2, 46)= 0.29, p=.75 |
| HC | [n=15] | - | 2.57 | 1.43 | |
| | | | (1.46) | (0.82) | |

**Supplementary Table 3d.** Mixed repeated measures ANOVAs (rmANOVA) across contingency ratings for each of the four conditions (ctx_unpred, ctx_safe, cue_pred, cue_safe) and each of the three phases (HAB, ACQ, EXT).

[**Abbreviations:** ACQ – Acquisition; CTX – Context; EXT – Extinction; HAB – Habituation; HC – Healthy control subjects without trauma experience; $p_{GG}$ – Greenhouse-Geisser correction; pred – Predictable; PTSD – patients with PTSD; SCR – Skin conductance response; TC – healthy control subjects with trauma experience; unpred – Unpredictable]

#### Ratings [Diff CS+-CS-]

| Arousal | | PTSD | TC | HC | Analyses |
|---|---|---|---|---|---|
| | | [n=19] | [n=17] | [n=19] | |
| HAB | | 1.06 (1.53) | 0.95 (1.41) | 0.76 (1.51) | F(2, 52)= 0.18, p=.83 |
| ACQ | Con | 0.21 (1.87) | 0.20 (1.62) | -0.45 (1.40) | Group: F(2, 56)= 0.64, p=.53<br>**Phase: F(1, 56)= 39.13, p<.001*****<br>$ACQ_{cue} > ACQ_{con}$ |
| | Cue | 1.97 (1.97) | 1.75 (2.51) | 1.48 (2.42) | GroupxPhase: F(2, 56)= 0.15, p=.86 |
| EXT | Con | 0.82 (1.67) | 0.15 (1.15) | 0.69 (1.62) | Group: F(2, 55)= 1.27, p=.29<br>Phase: F(1, 55)= 0.15, p=.70 |
| | Cue | 0.88 (1.47) | 0.30 (1.41) | 0.67 (0.94) | GroupxPhase: F(2, 55)= 0.11, p=.90 |
| **Valence** | | **PTSD** | **TC** | **HC** | **Analyses** |
| | | [n=19] | [n=17] | [n=19] | |
| HAB | | 0.85 (1.28) | 0.63 (1.63) | 0.26 (1.26) | F(2, 52)= 0.81, p=.45 |
| ACQ | Con | -0.13 (1.94) | 0.48 (1.60) | 0.48 (1.59) | Group: F(2, 56)= 1.11, p=.34<br>**Phase: F(1, 56)= 21.54, p<.001*****<br>$ACQ_{cue} > ACQ_{con}$ |
| | Cue | 1.71 (2.19) | 1.92 (2.12) | 1.25 (3.33) | GroupxPhase: F(2, 56)= 0.10, p=.90 |
| EXT | Con | 0.35 (1.52) | 0.60 (1.26) | 0.79 (1.52) | Group: F(2, 55)= 0.23, p=.80<br>Phase: F(1, 55)= 0.26, p=.61 |
| | Cue | 0.65 (1.61) | 0.95 (1.56) | 0.41 (1.06) | GroupxPhase: F(2, 55)= 1.98, p=.15 |
| **Contingency** | | **PTSD** | **TC** | **HC** | **Analyses** |
| | | [n=20] | [n=19] | [n=20] | |
| HAB | | 1.18 (2.21) | 1.08 (1.73) | 0.82 (2.43) | F(2, 52)= 0.14, p=.87 |
| ACQ | Con | -0.45 (2.19) | 0.00 (2.73) | -0.42 (1.42 | Group: F(2, 56)= 0.37, p=.69<br>**Phase: F(1, 56)= 42.89, p<.001*****<br>$ACQ_{cue} > ACQ_{con}$ |
| | Cue | 2.89 (3.55) | 3.25 (3.38) | 2.50 (3.39) | GroupxPhase: F(2, 56)= 0.07, p=.93 |
| EXT | Con | 0.68 (1.46) | 0.13 (1.44) | 0.57 (2.07) | Group: F(2, 55)= 0.70, p=.50<br>Phase: F(1, 55)= 0.37, p=.55 |
| | Cue | 1.00 (2.24) | 0.53 (1.37) | 0.26 (0.96) | GroupxPhase: F(2, 55)= 1.04, p=.36 |

**Suppl. Table 3e.** Difference of ratings between CS+ - CS- for the ratings of arousal**,** valence and contingency during all three phases (HAB, ACQ, EXT) and for all three groups (PTSD, TC, HC).

[**Abbreviations:** ACQ – Acquisition; Con – Context; CS – conditioned stimulus; EXT – Extinction; HAB – Habituation; HC – Healthy control subjects without trauma experience; PTSD – patients with PTSD; TC – healthy control subjects with trauma experience]

#### SCR (in μS)

| Groups | n | ctx unpred<br>M (SD) | ctx safe<br>M (SD) | Analyses | Cue pred<br>M (SD) | Cue pred<br>M (SD) | Analyses |
|---|---|---|---|---|---|---|---|
| **ACQ** | | | | | | | |
| PTSD | [n=14] | 0.011<br>(0.014) | 0.006<br>(0.010) | Group: F(2, 36)= 3.24, p=.051<br>**Context: F(1, 36)= 14.55,** | 0.028<br>(0.026) | 0.007<br>(0.009) | **Group: F(2, 34)= 5.45, p=.009****<br>PTSD < HC + TC |
| TC | [n=16] | 0.025<br>(0.022) | 0.017<br>(0.020) | **p<.001*****<br>ctx_unpred > ctx_safe | 0.052<br>(0.028) | 0.018<br>(0.015) | **Context: F(1, 34)= 66.07, p<.001*****<br>cue_pred > cue_safe |
| HC | [n=9] | 0.028<br>(0.019) | 0.020<br>(0.014) | Group x Context: F(2, 36)= 0.34,<br>p=.71 | 0.052<br>(0.009) | 0.023<br>(0.015) | Group x Context: F(2, 34)= 1.75,<br>p=.19 |
| **EXT** | | | | | | | |
| PTSD | [n=12] | 0.012<br>(0.017) | 0.010<br>(0.014) | Group: F(2, 35)= 1.78, p=.18<br>Context: F(1, 35)= 1.23, p=.28 | 0.009<br>(0.013) | 0.011<br>(0.014) | Group: F(2, 34)= 1.81, p=.18<br>Context: F(1, 34)= 2.31, p=.14 |
| TC | [n=16] | 0.015<br>(0.023) | 0.013<br>(0.024) | Group x Context: F(2, 35)= 0.04,<br>p=.96 | 0.015<br>(0.017) | 0.013<br>(0.018) | Group x Context: F(2, 34)= 2.25,<br>p=.12 |
| HC | [n=10] | 0.037<br>(0.058) | 0.035<br>(0.056) | | 0.040<br>(0.070) | 0.035<br>(0.065) | |

**Supplementary Table 4a.** Mixed repeated measures ANOVAs (rmANOVA) across SCRs for each of the two phases (context, cue) and each of the two phases (ACQ, EXT).

[**Abbreviations:** ACQ – Acquisition; CTX – Context; EXT – Extinction; HC – Healthy control subjects without trauma experience; pred – Predictable; PTSD – patients with PTSD; SCR – Skin conductance response; TC – healthy control subjects with trauma experience; unpred – Unpredictable; **μS** - Microsiemens]

### SCR (in µS)

**Diff. CS+-CS-**

| Groups | n | ctx unpred M (SD) | ctx safe M (SD) | Analyses | Cue pred M (SD) | Cue pred M (SD) | Analyses |
|---|---|---|---|---|---|---|---|
| **ACQ** | | | | | | | |
| PTSD | [n=14] | 0.003<br>(0.010) | 0.000<br>(0.006) | **Group: F(2, 36)= 3.93, p=.029***<br>HC > PTSD + TC | 0.059<br>(0.062) | 0.003<br>(0.007) | Group: F(2, 34)= 1.77, p=.19<br>**Context: F(1, 34)= 62.59, p<.001*** ** |
| TC | [n=16] | 0.004<br>(0.013) | -0.002<br>(0.009) | Context: F(1, 36)= 0.83, p=.37<br>Group x Context: F(2, 36)= 0.36, | 0.098<br>(0.046) | 0.004<br>(0.026) | cue_pred > cue_safe<br>Group x Context: F(2, 34)= 3.08, |
| HC | [n=9] | 0.008<br>(0.015) | 0.009<br>(0.019) | p=.70 | 0.066<br>(0.029) | 0.015<br>(0.023) | p=.059 |
| **EXT** | | | | | | | |
| PTSD | [n=12] | -0.017<br>(0.047) | 0.006<br>(0.016) | Group: F(2, 35)= 0.73, p=.49<br>**Context: F(1, 35)= 5.47, p=.025** | -0.001<br>(0.006) | 0.000<br>(0.011) | Group: F(2, 34)= 0.87, p=.43<br>Context: F(1, 34)= 1.57, p=.22 |
| TC | [n=16] | 0.001<br>(0.001) | 0.002<br>(0.012) | ctx_unpred_ext > ctx_safe_ext<br>Group x Context: F(2, 35)= 2.48, | 0.002<br>(0.009) | 0.001<br>(0.015) | Group x Context: F(2, 34)= 1.31,<br>p=.28 |
| HC | [n=10] | -0.017<br>(0.047) | 0.006<br>(0.016) | p=.098 | -0.022<br>(0.076) | 0.005<br>(0.014) | |

**Supplementary Table 4b.** Mixed repeated measures ANOVAs (rmANOVA) across SCRs for each of the two phases (context, cue) and each of the two phases (ACQ, EXT).

[**Abbreviations:** ACQ – Acquisition; CTX – Context; EXT – Extinction; HC – Healthy control subjects without trauma experience; pred – Predictable; PTSD – patients with PTSD; SCR – Skin conductance response; TC – healthy control subjects with trauma experience; unpred – Unpredictable; **µS** - Microsiemens]

| **Phase** | **Contexts** | **Contrast** | **k** | **Area of activation** | **MNI coordinates (x,y,z)** | **p(cluster-level)** |
|---|---|---|---|---|---|---|
| ACQ | ctx_unpred | HC>PTSD | 61 | Right middle cingulate gyrus | **9, -16, 29** | **.004*** |
| | | | 43 | Left thalamus | -6, -7, 2 | .014 |
| | | | 34 | Left parietal Operculum | -51, -25, 20 | .026 |
| | | | 31 | Right supramarginal Gyrus | 48, -34, 32 | .032 |
| | ctx_unpred > ctx_safectx | TC>PTSD | 37 | Right thalamus | 21, -19, 23 | .027 |
| | | | 29 | Right caudate/accumbens | 6, 11, -4 | .046 |
| EXT | ctx_pred | TC>PTSD | 35 | Right anterior cingulate gyrus | 12, 41, 5 | .050 |
| | ctx_unpred > ctx_safectx | PTSD>TC | 53 | Right caudate | 18, 23, 5 | .016 |
| | | | 53 | Left caudate | -9, 20, 5 | .016 |
| | | | 37 | Left hippocampus | -21, -40, 8 | .038 |

**Supplementary Table 5.** Whole brain contrasts for each of the two phases (ACQ, EXT) and between the three groups.

[**Abbreviations:** ACQ – Acquisition; CTX – Context; EXT – Extinction; HC – Healthy control subjects without trauma experience; MNI – coordinate system according to standard brains from the Montreal Neurological Institute; pred – Predictable; PTSD – patients with PTSD; TC – healthy control subjects with trauma experience; unpred – Unpredictable]

*cluster-level: p(FWE-corr.);

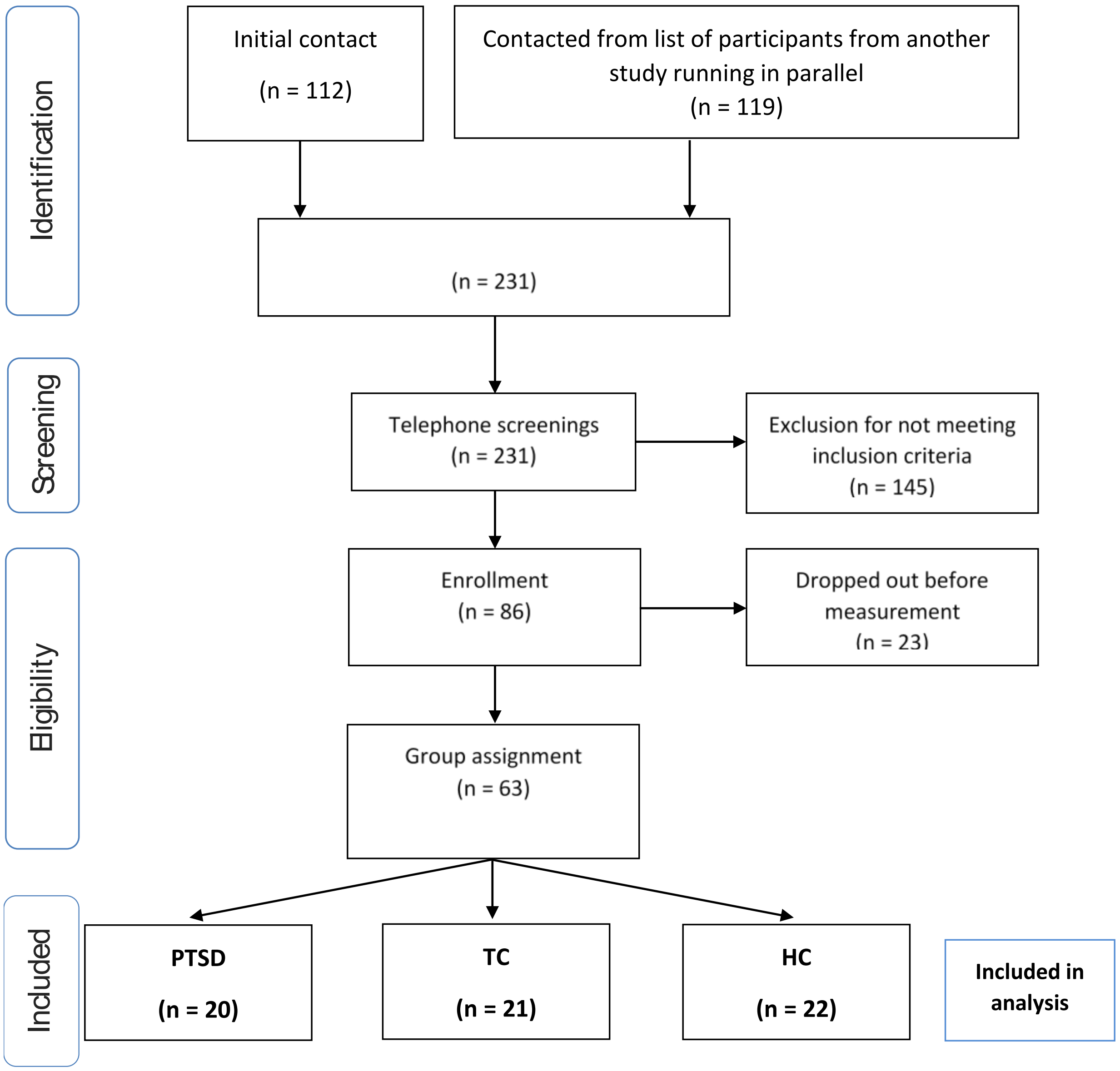


**Suppl. Figure 1.** Flowchart depicting identification, screening, eligibility and inclusion of subjects.

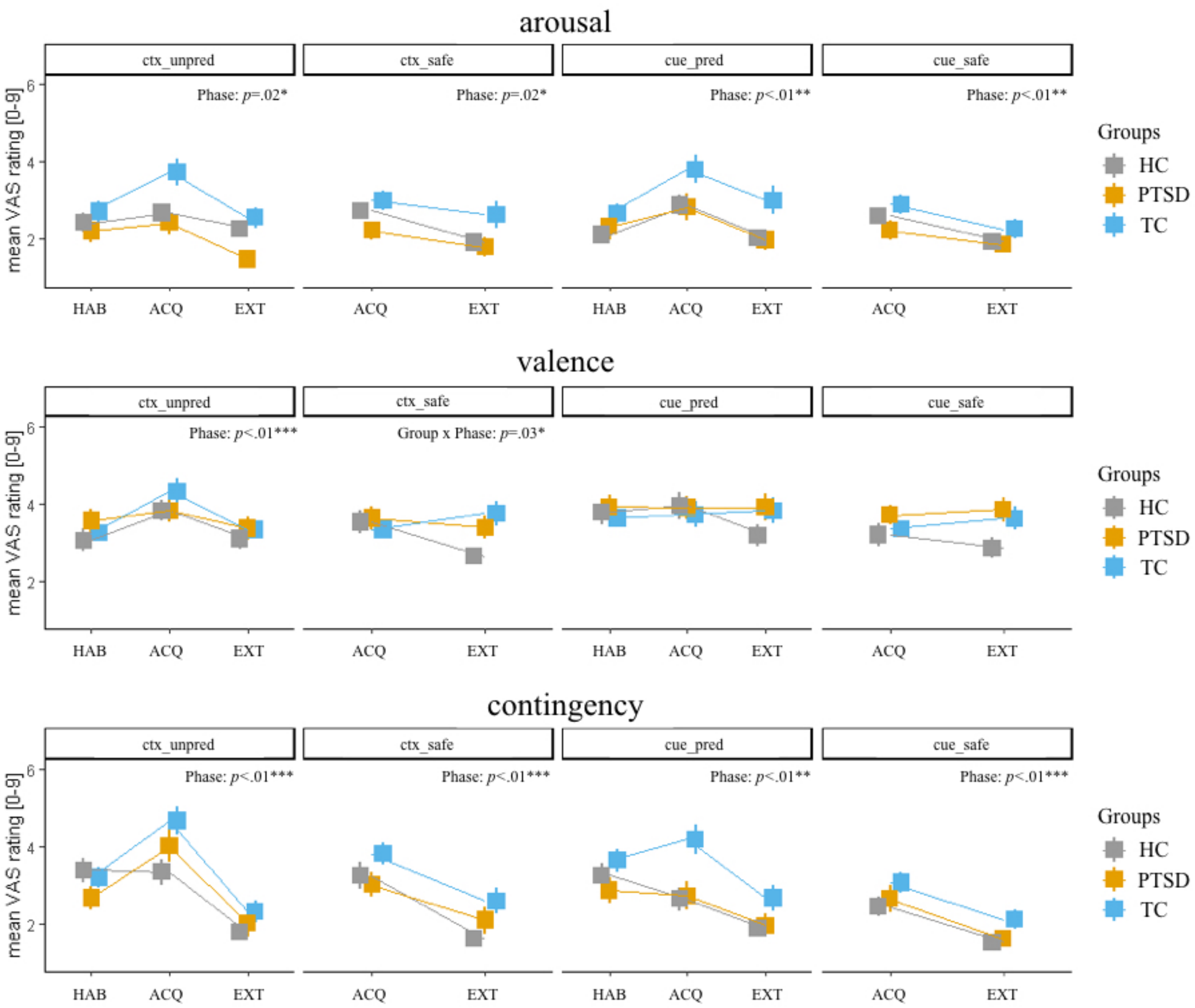

arousal
valence
contingency
ctx_unpred
ctx_safe
cue_pred
cue_safe
Phase: p=.02*
Phase: p=.02*
Phase: p<.01**
Phase: p<.01**
Phase: p<.01***
Group x Phase: p=.03*
Phase: p<.01***
Phase: p<.01***
Phase: p<.01**
Phase: p<.01***
mean VAS rating [0-9]
HAB
ACQ
EXT
Groups
HC
PTSD
TC

**Suppl. Figure 2a.** Arousal, valence and contingency ratings across each of the four conditions (ctx_unpred, ctx_safe, cue_pred, cue_safe), each of the three phases (HAB, ACQ, EXT) and each group (HC, PTSD, TC).

[**Abbreviations:** ACQ – Acquisition; CTX – Context; EXT – Extinction; HAB – Habituation; HC – Healthy control subjects without trauma experience; $p_{GG}$ – Greenhouse-Geisser correction; pred – Predictable; PTSD – patients with PTSD; SCR – Skin conductance response; TC – healthy control subjects with trauma experience; unpred – Unpredictable; VAS – Visual Analogue Scale]

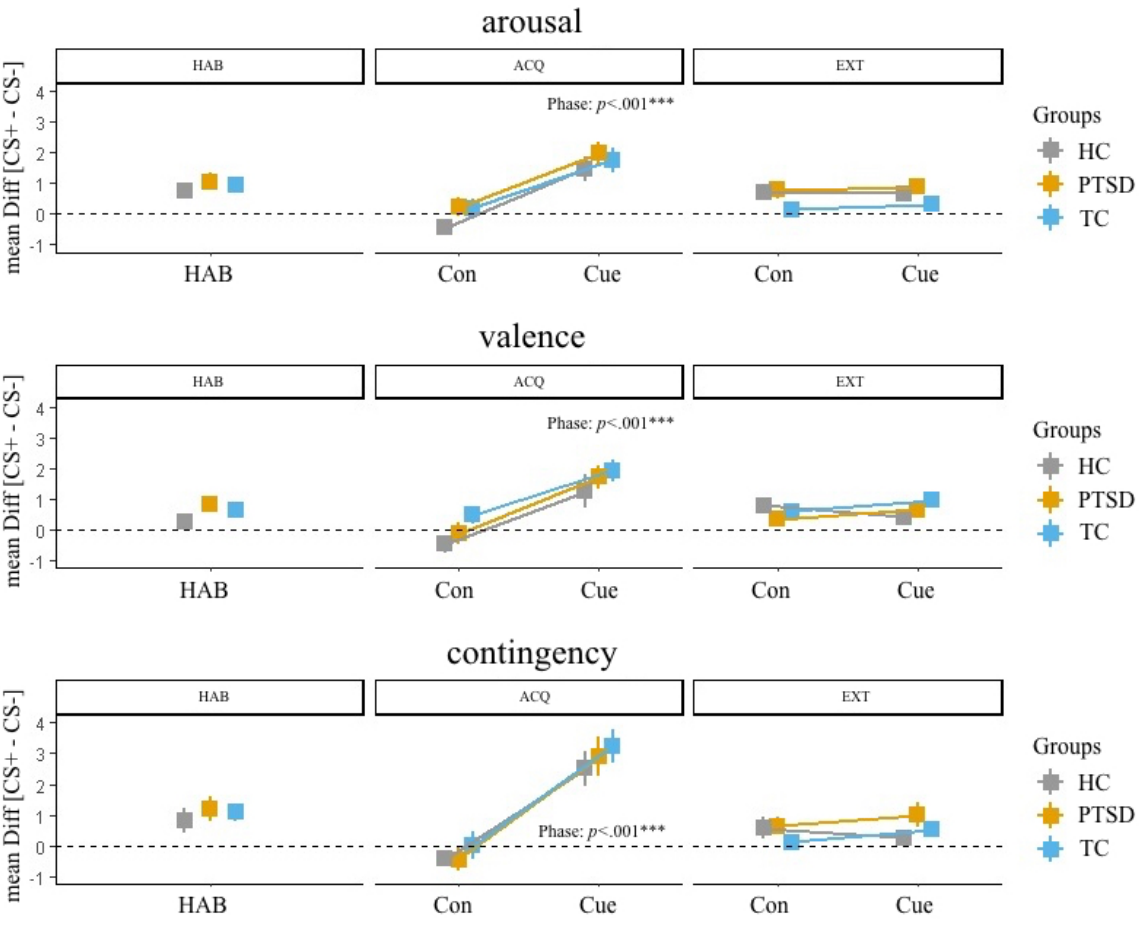


**Suppl. Figure 2b.** Difference scores (CS+ - CS-) for arousal, valence and contingency ratings across each of the three phases (HAB, ACQ, EXT) and each group (HC, PTSD, TC).

[**Abbreviations:** ACQ – Acquisition; Con – Context; EXT – Extinction; HAB – Habituation; HC – Healthy control subjects without trauma experience; PTSD – patients with PTSD; TC – healthy control subjects with trauma experience]

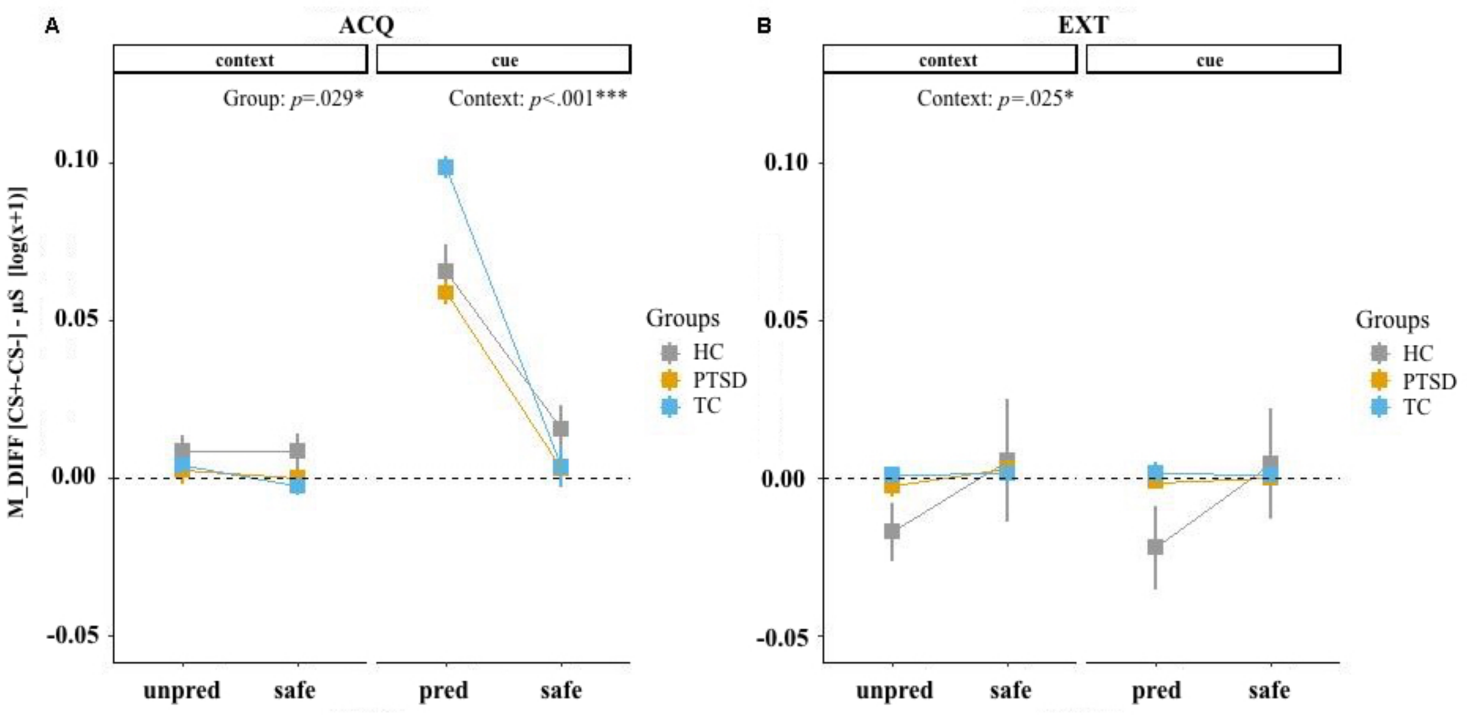


**Suppl. Figure 3.** Difference scores of CS+-CS- of the SCRs across each of the four conditions (CTX_unpred, CTX_safe, CUE_pred, CUE_safe), two phases (ACQ, EXT) and each group (HC, PTSD, TC). A) ACQ phase. B) EXT phase.

[**Abbreviations:** ACQ – Acquisition; CTX – Context; EXT – Extinction; HC – Healthy control subjects without trauma experience; M_DIFF – Mean difference of CS+ - CS- SCR; pred – Predictable; PTSD – patients with PTSD; SCR – Skin conductance response; TC – healthy control subjects with trauma experience; unpred – Unpredictable]
