## Supplementary material for "Altered frontolimbic activity during virtual reality-based contextual fear learning in patients with posttraumatic stress disorder": Suppl. Fig 1

**Suppl. Figure 1.** Flow Diagram

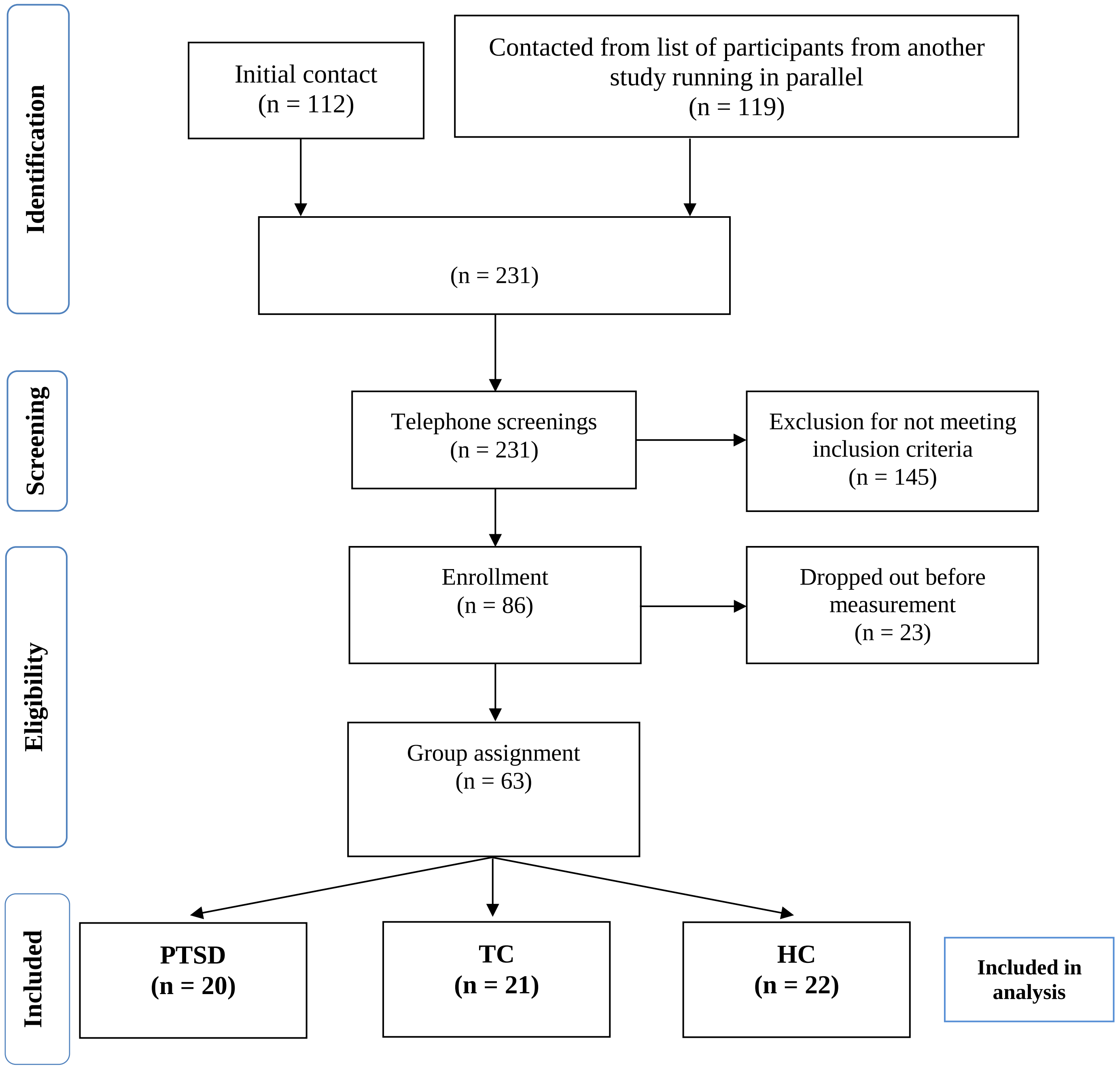


**Suppl. Figure 1.** Flowchart depicting identification, screening, eligibility and inclusion of subjects.
