## Supplementary material for "Altered frontolimbic activity during virtual reality-based contextual fear learning in patients with posttraumatic stress disorder": Suppl. Fig 2

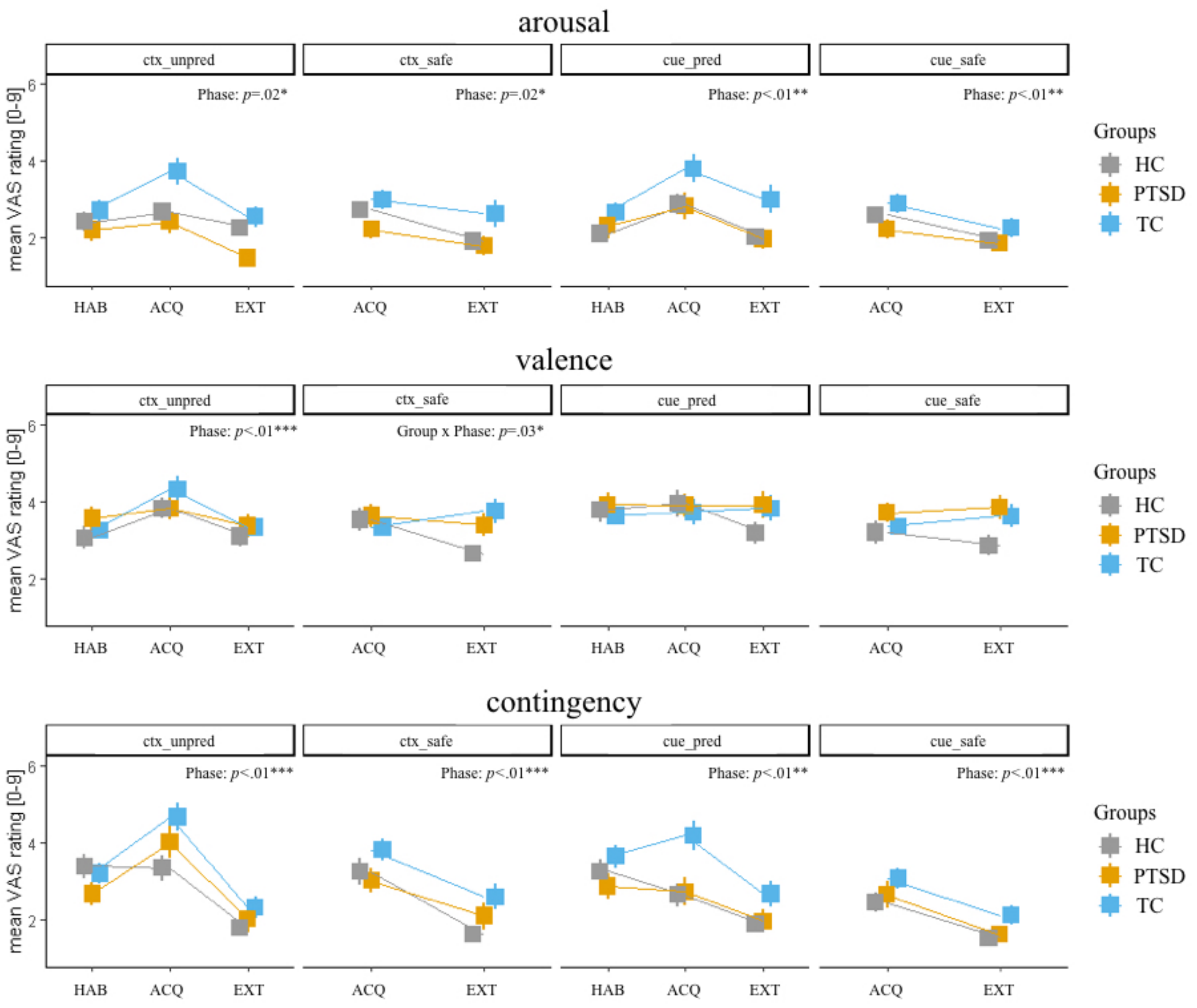
arousal
ctx_unpred
ctx_safe
cue_pred
cue_safe
Phase: p=.02*
Phase: p=.02*
Phase: p<.01**
Phase: p<.01**
mean VAS rating [0-9]
HAB
ACQ
EXT
Groups
HC
PTSD
TC
valence
Phase: p<.01***
Group x Phase: p=.03*
contingency
Phase: p<.01***
Phase: p<.01***
Phase: p<.01**
Phase: p<.01***

**Suppl. Figure 2a.** Arousal, valence and contingency ratings across each of the four conditions (ctx_unpred, ctx_safe, cue_pred, cue_safe), each of the three phases (HAB, ACQ, EXT) and each group (HC, PTSD, TC).

[**Abbreviations:** ACQ – Acquisition; CTX – Context; EXT – Extinction; HAB – Habituation; HC – Healthy control subjects without trauma experience; $p_{GG}$ – Greenhouse-Geisser correction; pred – Predictable; PTSD – patients with PTSD; SCR – Skin conductance response; TC – healthy control subjects with trauma experience; unpred – Unpredictable; VAS – Visual Analogue Scale]

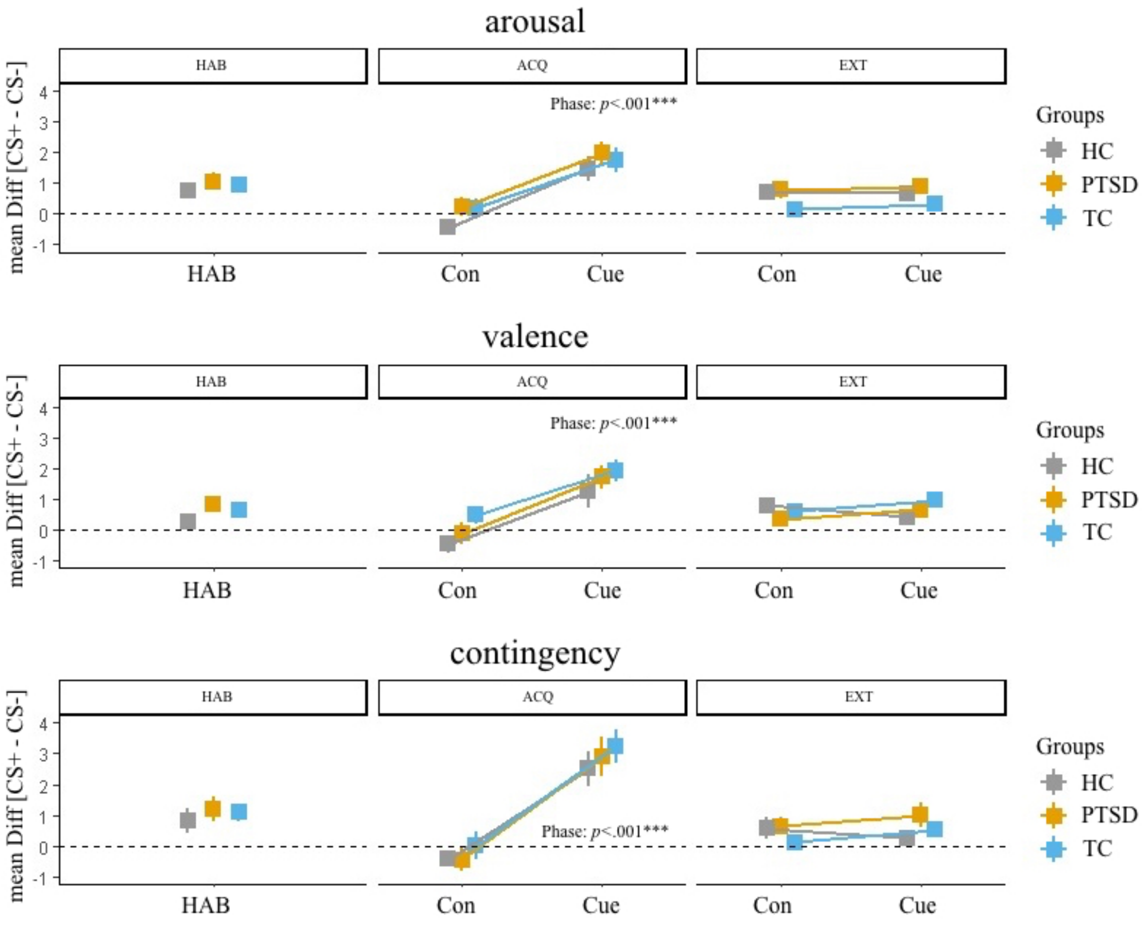


**Suppl. Figure 2b.** Difference scores (CS+ - CS-) for arousal, valence and contingency ratings across each of the three phases (HAB, ACQ, EXT) and each group (HC, PTSD, TC).

[**Abbreviations:** ACQ – Acquisition; Con – Context; EXT – Extinction; HAB – Habituation; HC – Healthy control subjects without trauma experience; PTSD – patients with PTSD; TC – healthy control subjects with trauma experience]
