## Supplementary material for "Altered frontolimbic activity during virtual reality-based contextual fear learning in patients with posttraumatic stress disorder": Suppl. Fig 3

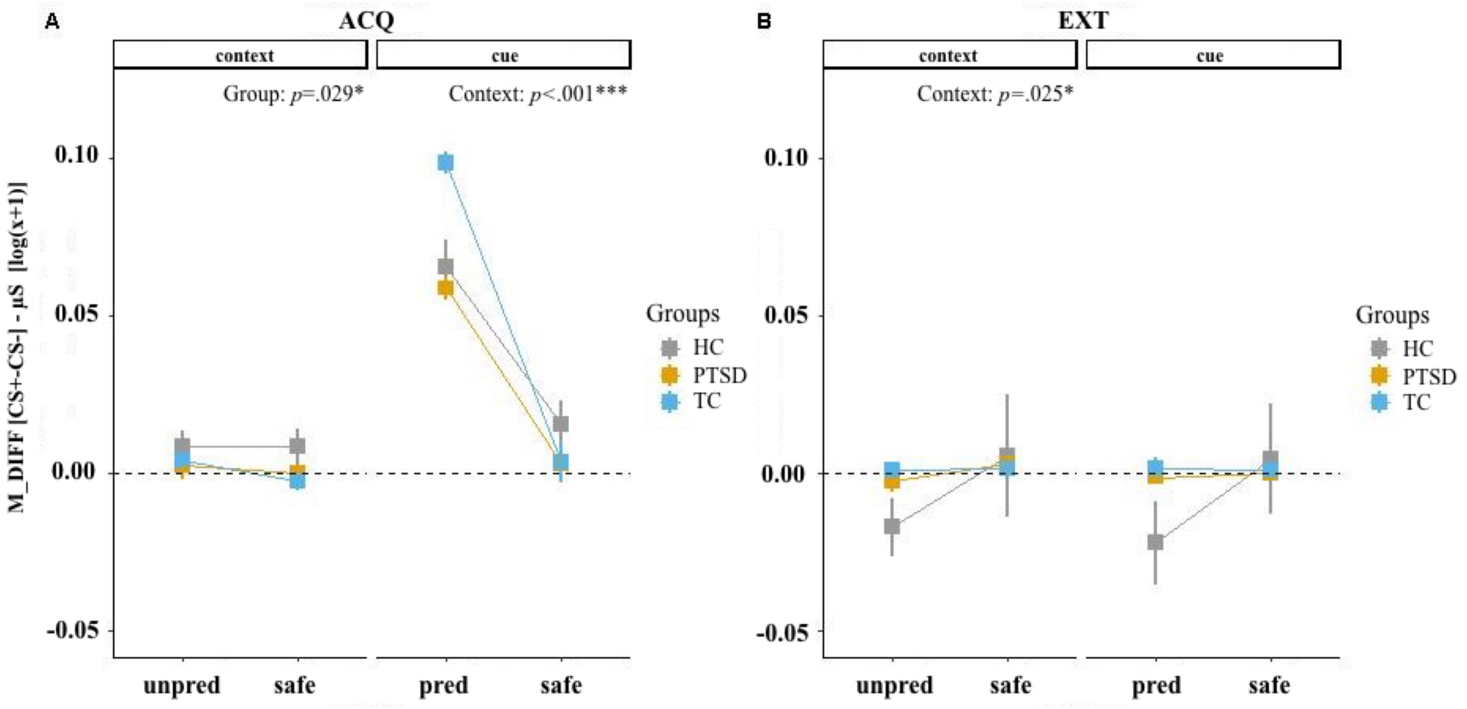


**Suppl. Figure 3.** Difference scores of CS+-CS- of the SCRs across each of the four conditions (CTX_unpred, CTX_safe, CUE_pred, CUE_safe), two phases (ACQ, EXT) and each group (HC, PTSD, TC). A) ACQ phase. B) EXT phase.

[**Abbreviations:** ACQ – Acquisition; CTX – Context; EXT – Extinction; HC – Healthy control subjects without trauma experience; M_DIFF – Mean difference of CS+ - CS- SCR; pred – Predictable; PTSD – patients with PTSD; SCR – Skin conductance response; TC – healthy control subjects with trauma experience; unpred – Unpredictable]
