## Supplementary material for "Altered frontolimbic activity during virtual reality-based contextual fear learning in patients with posttraumatic stress disorder": Suppl. Tab 3

## US

| Groups | n | HAB<br>M (SD) | ACQ Con<br>M (SD) | ACQ Cue<br>M (SD) | Analyses |
|---|---|---|---|---|---|
| **Intensity (in mA)** | | | | | |
| PTSD | [n=19] | 4.68 (3.39) | - | - | F(2, 58)= 0.54, p=.59 |
| TC | [n=20] | 4.80 (2.41) | - | - | |
| HC | [n=22] | 3.96 (2.72) | - | - | |
| **Pain** | | | | | |
| PTSD | [n=17] | 7.29 (0.77) | 5.47 (1.94) | 5.94 (1.71) | Group: F(2, 54)= 2.29, p=.11<br>**Phase: F(2, 108)= 31.27, p<.001*****<br>HAB > $ACQ_{Con}$ + $ACQ_{Cue}$ |
| TC | [n=20] | 7.10 (0.45) | 6.55 (0.76) | 6.55 (0.76) | **GroupxPhase: F(4, 108)= 3.05, p=.02***<br>$TC_{ACQ\_con}$ > $PTSD_{ACQ\_con}$ + $HC_{ACQ\_con}$ |
| HC | [n=20] | 7.25 (0.44) | 5.40 (2.06) | 5.50 (1.82) | $TC_{ACQ\_cue}$ > $PTSD_{ACQ\_cue}$ + $HC_{ACQ\_cue}$ |
| **Valence** | | | | | |
| PTSD | [n=17] | 7.29 (0.69) | 5.76 (1.92) | 6.18 (1.88) | Group: F(2, 54)= 3.02, p=.057<br>**Phase: F(2, 108)= 18.46, $p_{GG}$<.001*****<br>HAB > $ACQ_{Con}$ + $ACQ_{Cue}$ |
| TC | [n=20] | 7.10 (0.45) | 6.75 (1.02) | 6.85 (1.18) | **GroupxPhase: F(4, 108)= 3.06, $p_{GG}$=.031***<br>$TC_{ACQ\_con}$ > $PTSD_{ACQ\_con}$ + $HC_{ACQ\_con}$ |
| HC | [n=20] | 7.10 (0.55) | 5.60 (1.90) | 5.35 (2.21) | $TC_{ACQ\_cue}$ > $HC_{ACQ\_cue}$ |

**Suppl. Table 3a.** Intensity (in Milliampere), pain intensity ratings and valence ratings of the US during HAB and ACQ.

[**Abbreviations:** ACQ – Acquisition; Con – Context; EXT – Extinction; HAB – Habituation; HC – Healthy control subjects without trauma experience; mA – Milliampere; PTSD – patients with PTSD; TC – healthy control subjects with trauma experience; US – Unconditioned Stimulus]

#### Arousal

| Groups | n | HAB M (SD) | ACQ M (SD) | EXT M (SD) | Analyses |
|---|---|---|---|---|---|
| **CTX_unpred** | | | | | |
| PTSD | [n=14] | 2.21 (1.73) | 2.46 (1.68) | 1.54 (0.91) | Group: F(2, 41)= 1.40, p=.26<br>**Phase: F(2, 82)= 4.33, $p_{GG}$=.022***<br>ACQ > HAB + EXT<br>GroupxPhase: F(4, 82)= 1.38, p=.25 |
| TC | [n=17] | 2.41 (1.24) | 3.47 (2.22) | 2.24 (1.44) | |
| HC | [n=13] | 2.15 (1.25) | 2.27 (0.90) | 2.23 (1.13) | |
| **CTX_safe** | | | | | |
| PTSD | [n=16] | - | 2.31 (1.52) | 1.69 (1.40) | Group: F(2, 46)= 0.80, p=.46<br>**Phase: F(1, 46)= 6.45, p=.015***<br>ACQ > EXT<br>GroupxPhase: F(2, 46)= 0.33, p=.72 |
| TC | [n=18] | - | 2.72 (1.22) | 2.42 (2.10) | |
| HC | [n=15] | - | 2.60 (1.45) | 1.90 (1.14) | |
| **CUE_pred** | | | | | |
| PTSD | [n=14] | 2.46 (1.83) | 2.83 (1.83) | 2.00 (1.40) | Group: F(2, 42)= 1.30, p=.28<br>**Phase: F(2, 82)= 6.03, p=.004****<br>ACQ > HAB + EXT<br>GroupxPhase: F(4, 82)= 0.56, p=.69 |
| TC | [n=17] | 2.50 (1.35) | 3.62 (2.33) | 2.79 (2.27) | |
| HC | [n=13] | 1.85 (0.90) | 2.62 (1.34) | 2.04 (1.25) | |
| **CUE_safe** | | | | | |
| PTSD | [n=16] | - | 2.28 (1.48) | 1.78 (1.03) | Group: F(2, 46)= 0.34, p=.72<br>**Phase: $F_P$(1, 46)= 9.55, p=.003****<br>ACQ > EXT<br>GroupxPhase: F(2, 46)= 0.12, p=.89 |
| TC | [n=18] | - | 2.56 (1.12) | 2.08 (1.41) | |
| HC | [n=15] | - | 2.63 (1.25) | 1.97 (1.34) | |

**Supplementary Table 3b.** Mixed repeated measures ANOVAs (rmANOVA) across arousal ratings for each of the four conditions (ctx_unpred, ctx_safe, cue_pred, cue_safe) and each of the three phases (HAB, ACQ, EXT).

[**Abbreviations:** ACQ – Acquisition; CTX – Context; EXT – Extinction; HAB – Habituation; HC – Healthy control subjects without trauma experience; $p_{GG}$ – Greenhouse-Geisser correction; pred – Predictable; PTSD – patients with PTSD; SCR – Skin conductance response; TC – healthy control subjects with trauma experience; unpred – Unpredictable]

### Valence

| Groups | n | HAB M (SD) | ACQ M (SD) | EXT M (SD) | Analyses |
|---|---|---|---|---|---|
| **CTX_unpred** | | | | | |
| PTSD | [n=14] | 3.54 (1.67) | 3.71 (1.99) | 3.36 (1.79) | Group: $F(2, 41)= 0.30, p=.74$<br>**Phase: $F(2, 82)= 8.90, p_{GG}<.001$*****<br>ACQ > HAB + EXT<br>GroupxPhase: $F(4, 82)= 0.92, p=.45$ |
| TC | [n=17] | 3.26 (1.38) | 4.18 (2.08) | 3.18 (1.49) | |
| HC | [n=13] | 2.81 (1.49) | 3.85 (1.63) | 2.81 (1.63) | |
| **CTX_safe** | | | | | |
| PTSD | [n=16] | - | 3.53 (1.79) | 3.22 (1.81) | Group: $F(2, 46)= 0.86, p=.43$<br>Phase: $F(1, 46)= 2.26, p=.14$<br>**GroupxPhase: $F(2, 46)= 3.69, p=.033$***<br>$HC_{EXT} > PTSD_{EXT} + TC_{EXT}$ |
| TC | [n=18] | - | 3.33 (1.37) | 3.72 (2.12) | |
| HC | [n=15] | - | 3.40 (1.66) | 2.33 (0.96) | |
| **CUE_pred** | | | | | |
| PTSD | [n=14] | 3.89 (1.91) | 3.63 (1.91) | 4.00 (2.12) | Group: $F(2, 42)= 0.19, p=.83$<br>Phase: $F(2, 82)= 0.19, p=.83$<br>GroupxPhase: $F(4, 82)= 1.08, p=.37$ |
| TC | [n=17] | 3.65 (1.43) | 3.65 (1.94) | 3.88 (2.18) | |
| HC | [n=13] | 3.37 (1.91) | 3.81 (1.68) | 2.96 (1.89) | |
| **CUE_safe** | | | | | |
| PTSD | [n=16] | - | 3.69 (1.71) | 3.84 (1.94) | Group: $F(2, 46)= 1.83, p=.17$<br>Phase: $F_P(1, 46)= 0.01, p=.94$<br>GroupxPhase: $F(2, 46)= 1.29, p=.29$ |
| TC | [n=18] | - | 3.25 (1.31) | 3.61 (1.92) | |
| HC | [n=15] | - | 3.00 (1.21) | 2.53 (1.70) | |

**Supplementary Table 3c.** Mixed repeated measures ANOVAs (rmANOVA) across valence ratings for each of the four conditions (ctx_unpred, ctx_safe, cue_pred, cue_safe) and each of the three phases (HAB, ACQ, EXT).

[**Abbreviations:** ACQ – Acquisition; CTX – Context; EXT – Extinction; HAB – Habituation; HC – Healthy control subjects without trauma experience; $p_{GG}$ – Greenhouse-Geisser correction; pred – Predictable; PTSD – patients with PTSD; SCR – Skin conductance response; TC – healthy control subjects with trauma experience; unpred – Unpredictable]

#### Contingency

| Groups | n | HAB M (SD) | ACQ M (SD) | EXT M (SD) | Analyses |
|---|---|---|---|---|---|
| **CTX_unpred** | | | | | |
| PTSD | [n=14] | 2.64 (1.70) | 3.68 (1.87) | 2.11 (2.26) | Group: $F(2, 41)= 1.12$, $p=.34$<br>**Phase: $F(2, 82)= 10.56$, $p_{GG}<.001$*****<br>EXT < HAB + ACQ<br>GroupxPhase: $F(4, 82)= 0.55$, $p=.70$ |
| TC | [n=17] | 3.12 (1.60) | 4.29 (1.98) | 2.38 (1.75) | |
| HC | [n=13] | 3.08 (1.80) | 3.12 (2.58) | 1.58 (1.10) | |
| **CTX_safe** | | | | | |
| PTSD | [n=16] | - | 3.22 (1.91) | 2.06 (2.15) | Group: $F(2, 46)= 0.92$, $p=.41$<br>**Phase: $F(1, 46)= 13.36$, $p<.001$*****<br>ACQ > EXT<br>GroupxPhase: $F(2, 46)= 0.42$, $p=.66$ |
| TC | [n=18] | - | 3.64 (1.75) | 2.64 (2.17) | |
| HC | [n=15] | - | 3.33 (2.53) | 1.57 (0.89) | |
| **CUE_pred** | | | | | |
| PTSD | [n=14] | 2.89 (1.96) | 2.75 (2.03) | 1.89 (1.71) | Group: $F(2, 42)= 2.65$, $p=.83$<br>**Phase: $F(2, 82)= 5.27$, $p=.007$****<br>EXT < HAB + ACQ<br>GroupxPhase: $F(4, 82)= 0.53$, $p=.72$ |
| TC | [n=17] | 3.41 (1.65) | 4.24 (2.22) | 2.79 (2.28) | |
| HC | [n=13] | 2.81 (1.74) | 2.62 (1.96) | 1.85 (1.39) | |
| **CUE_safe** | | | | | |
| PTSD | [n=16] | - | 2.78 (1.83) | 1.56 (1.14) | Group: $F(2, 46)= 0.99$, $p=.38$<br>**Phase: $F_P(1, 46)= 16.57$, $p<.001$*****<br>ACQ > EXT<br>GroupxPhase: $F(2, 46)= 0.29$, $p=.75$ |
| TC | [n=18] | - | 2.94 (1.70) | 2.17 (1.56) | |
| HC | [n=15] | - | 2.57 (1.46) | 1.43 (0.82) | |

**Supplementary Table 3d.** Mixed repeated measures ANOVAs (rmANOVA) across contingency ratings for each of the four conditions (ctx_unpred, ctx_safe, cue_pred, cue_safe) and each of the three phases (HAB, ACQ, EXT).

[**Abbreviations:** ACQ – Acquisition; CTX – Context; EXT – Extinction; HAB – Habituation; HC – Healthy control subjects without trauma experience; $p_{GG}$ – Greenhouse-Geisser correction; pred – Predictable; PTSD – patients with PTSD; SCR – Skin conductance response; TC – healthy control subjects with trauma experience; unpred – Unpredictable]

**Ratings [Diff CS+-CS-]**

| **Arousal** | | **PTSD** | **TC** | **HC** | **Analyses** |
|---|---|---|---|---|---|
| | | [n=19] | [n=17] | [n=19] | |
| HAB | | 1.06 | 0.95 | 0.76 | F(2, 52)= 0.18, p=.83 |
| | | (1.53) | (1.41) | (1.51) | |
| ACQ | Con | 0.21 | 0.20 | -0.45 | Group: F(2, 56)= 0.64, p=.53 |
| | | (1.87) | (1.62) | (1.40) | **Phase: F(1, 56)= 39.13, p<.001*** ** |
| | Cue | 1.97 | 1.75 | 1.48 | $ACQ_{cue} > ACQ_{con}$ |
| | | (1.97) | (2.51) | (2.42) | GroupxPhase: F(2, 56)= 0.15, p=.86 |
| EXT | Con | 0.82 | 0.15 | 0.69 | Group: F(2, 55)= 1.27, p=.29 |
| | | (1.67) | (1.15) | (1.62) | Phase: F(1, 55)= 0.15, p=.70 |
| | Cue | 0.88 | 0.30 | 0.67 | GroupxPhase: F(2, 55)= 0.11, p=.90 |
| | | (1.47) | (1.41) | (0.94) | |
| **Valence** | | **PTSD** | **TC** | **HC** | **Analyses** |
| | | [n=19] | [n=17] | [n=19] | |
| HAB | | 0.85 | 0.63 | 0.26 | F(2, 52)= 0.81, p=.45 |
| | | (1.28) | (1.63) | (1.26) | |
| ACQ | Con | -0.13 | 0.48 | 0.48 | Group: F(2, 56)= 1.11, p=.34 |
| | | (1.94) | (1.60) | (1.59) | **Phase: F(1, 56)= 21.54, p<.001*** ** |
| | Cue | 1.71 | 1.92 | 1.25 | $ACQ_{cue} > ACQ_{con}$ |
| | | (2.19) | (2.12) | (3.33) | GroupxPhase: F(2, 56)= 0.10, p=.90 |
| EXT | Con | 0.35 | 0.60 | 0.79 | Group: F(2, 55)= 0.23, p=.80 |
| | | (1.52) | (1.26) | (1.52) | Phase: F(1, 55)= 0.26, p=.61 |
| | Cue | 0.65 | 0.95 | 0.41 | GroupxPhase: F(2, 55)= 1.98, p=.15 |
| | | (1.61) | (1.56) | (1.06) | |
| **Contingency** | | **PTSD** | **TC** | **HC** | **Analyses** |
| | | [n=20] | [n=19] | [n=20] | |
| HAB | | 1.18 | 1.08 | 0.82 | F(2, 52)= 0.14, p=.87 |
| | | (2.21) | (1.73) | (2.43) | |
| ACQ | Con | -0.45 | 0.00 | -0.42 | Group: F(2, 56)= 0.37, p=.69 |
| | | (2.19) | (2.73) | (1.42 | **Phase: F(1, 56)= 42.89, p<.001*** ** |
| | Cue | 2.89 | 3.25 | 2.50 | $ACQ_{cue} > ACQ_{con}$ |
| | | (3.55) | (3.38) | (3.39) | GroupxPhase: F(2, 56)= 0.07, p=.93 |
| EXT | Con | 0.68 | 0.13 | 0.57 | Group: F(2, 55)= 0.70, p=.50 |
| | | (1.46) | (1.44) | (2.07) | Phase: F(1, 55)= 0.37, p=.55 |
| | Cue | 1.00 | 0.53 | 0.26 | GroupxPhase: F(2, 55)= 1.04, p=.36 |
| | | (2.24) | (1.37) | (0.96) | |

**Suppl. Table 3e.** Difference of ratings between CS+ - CS- for the ratings of arousal**,** valence and contingency during all three phases (HAB, ACQ, EXT) and for all three groups (PTSD, TC, HC).

[**Abbreviations:** ACQ – Acquisition; Con – Context; CS – conditioned stimulus; EXT – Extinction; HAB – Habituation; HC – Healthy control subjects without trauma experience; PTSD – patients with PTSD; TC – healthy control subjects with trauma experience]
